# A Formal Model for the Representation of Binary Temporal Relations in Healthcare Applications and an Efficient Algorithm for Logic-Based Temporal Subsumption Testing and Pattern Matching

**DOI:** 10.1101/2023.11.17.23298715

**Authors:** Walter Sujansky, Keith E. Campbell

## Abstract

**Objectives:** Important temporal relationships exist among pairs of medically relevant events stored in electronic health records (EHRs), such as “the infection began within two weeks after surgery”. Queries for specific temporal patterns also appear in decision-support rules and data-analysis programs. The accurate matching of such patterns to the patient data in EHRs is critical to the effective performance of decision-support systems, statistical analysis programs, data-abstraction processes, digital phenotyping for machine-learning, and other applications. The correct classification of temporally-qualified concepts in biomedical terminologies and ontologies, such as SNOMED-CT, is also important to ensure the accuracy and completeness of these knowledge-based resources.

**Methods:** In this paper, we describe an expressive model to formally represent temporal relationships between pairs of events, including “Before”, “During”, “Within *n* days after”, and “Within *n* hours before or *m* hours after, but not during”. We also describe a novel logic-based algorithm to deduce whether one such relationship temporally matches (i.e., is subsumed by) another such relationship, which enables the querying of structured time-stamped patient data, the querying of semi-structured narrative patient data, and the classification of logically defined medical concepts. Our model assumes an interval-based notion of time and our algorithm implements a logic-based definition of subsumption.

**Results:** We formally prove the correctness of the algorithm based on the properties of temporal intervals and the axioms of propositional logic. We also prove that the algorithm has computational complexity of constant-time (i.e., O(1)) with respect to the size of the database being queried or the knowledge base being classified.

**Conclusion:** The novel model and algorithm described here for temporal representation and reasoning are sound and have the potential to facilitate temporal subsumption testing and pattern matching in a number of medical application domains. Empirical testing is needed to establish the full scope of useful applicability.

## 1 Introduction and Background

Temporal relationships between pairs of medically relevant events are often critical in determining the causes, significance and treatment of clinical disorders. For example, a cardiac arrhythmia occurring during surgery may be investigated and managed very differently than one occurring two months after surgery, given the potential role of anesthetic agents in the former. Similarly, a serious maternal infection during the first trimester of pregnancy may have different implications and warrant different management than one occurring during the third trimester. The representation of such temporal relationships, therefore, is an important aspect of modeling both patient data and medical knowledge in clinical information systems.

In addition, the automated application of medical knowledge to patient data, as well as the sophisticated analysis of patient data, require robust and efficient algorithms to match temporal relationships recorded in patient data with temporal patterns specified in decision-support rules, in statistical abstraction categories, or in machine-learning parameters. For example, a practice guideline for managing infectious disease may specify that certain blood cultures should be ordered for any infection that occurs within two weeks following a surgery, while a patient’s timestamped record may document that a skin infection began on Jan. 16^th^, 2023 and the patient had an appendectomy on Jan. 10^th^, 2023. Alternatively, the patient’s record may simply include the physician’s narrative comment “the patient had an appendectomy 6 days prior to first noticing the infection.” For an intelligent system to automatically apply the logic of the practice guideline to such data, it must have the capability to determine whether the relative timing of the patient’s infection and surgery, represented in either of these ways, satisfies the temporal pattern specified in the practice guideline. In general, this capability requires a model for representing relevant temporal relationships in a manner that is expressive and that supports a powerful and reliable pattern-matching and subsumption-testing algorithm.

### 1.1 Previous Work

Substantial past work has explored the representation of temporal relationships between pairs of events and reasoning over such relationships, such as the matching of temporal relationship to time-stamped data or the comparison of temporal relationships to each other. Allen’s seminal work [1] identified a complete set of 13 qualitative relations between pairs of temporal intervals, such as “precedes”,

“during”, and “overlaps”, and specified a calculus for inferring new relations based on asserted temporal facts. Although influential in subsequent research, Allen’s model did not support the representation of quantitative (metric) relationships between temporal intervals (such as “Interval A precedes Interval B by at least 24 hours, but by less than 1 week”), nor did it propose any mechanisms for evaluating temporal intervals with respect to instance data (such as “find all patients in whom a vaccination preceded a severe allergic reaction by at least 24 hours, but by less than 1 week”).

Subsequent researchers [2] modeled metric temporal relationships between time points as linear inequalities of the form “t_2_ – t_1_ *≥* n”, which denotes that timepoint 1 precedes timepoint 2 by at least *n* time units. Conjunctions of such linear inequalities allowed the representation temporal relationships that include *ranges*, such as “t_2_ – t_1_ *≥* 24 hours ^ t_2_ – t_1_ < 1 week. Permitting the values 0 and ∞ as constants in such expressions further extended their expressiveness, allowing the metric representation of quantitative or qualitative relationships (such as Allen’s original qualitative relations) [3,4]. For example, the semantics of the Allen interval relation “Interval 1 precedes Interval 2” could be represented as the conjunction “b_2_ – e_1_ > 0 ^ b_2_ – e_1_ < ∞”, where “b_2_” denotes the beginning time point of Interval 2 and “e_1_” denotes the ending time point of Interval 1.

Research on automated reasoning over such metric representations primarily addressed two tasks. First, as with Allen’s work, algorithms were developed to compute the implicit temporal relationships inferred by a set of asserted linear inequalities, i.e. temporal constraint propagation [5]. For example, from the assertions “Given that Dick leaves for work within 10 minutes of 8:00 AM and his commute takes between 30 and 45 minutes, and Jane arrives at work within 15 minutes of 9:00 AM, is it possible for Jane to arrive at work before Dick? What is the latest after Dick that Jane could arrive?” Such tasks were useful for AI planning applications. The second task entailed *solving* a set of linear temporal inequalities, i.e., computing the set of time points that collectively satisfy all of the constraints represented by the inequalities [2]. This task, often characterized as the Temporal Constraint Satisfaction Problem (TCSP), addressed a variety of automated planning and scheduling problems.

However, fewer researchers addressed methods to match temporal relationships formally expressed as combinations of linear inequalities to time-stamped or temporally-annotated data. This “temporal pattern matching” (TPM) task aims to identify all existing data instances that satisfy a set of temporal constraints or to evaluate whether a specific data instance satisfies such a set. Many applications of TMP exist in the health care domain.

TMP is an important task in identifying cohorts of patients in clinical data repositories that satisfy the eligibility criteria for randomized trials or observational research studies, which can often be complex [6, 7,8]. For example, a study may only include patients who experienced a certain condition within one week after being administered a suspected medication, while excluding patients who were taking more than two other medications at the time the condition occurred or who had previously experienced the condition in the six months prior to being administered the suspected medication. TMP is also important in correctly applying automated clinical decision support rules to patient data, including clinical practice guidelines and alerts, when the logic of such rules specifies a particular sequence of two events or a specific temporal proximity of two events [9]. For example, an alert rule may be programmed to suggest the administration of a COVID vaccine only for patients with no positive COVID test during the 90 days prior to their visit. The specifications for electronic clinical quality measures also include temporal inclusion and exclusion criteria for correctly computing whether health care providers performed tests or interventions associated with high-quality care [10]. Beyond these data-analytical needs, the logical definitions of certain medical concepts in clinical terminologies, such as SNOMED-CT, also include temporal qualifiers. For example, a “post-surgical infection” is formally defined as the concept “infection” occurring after the concept “surgery”. The correct automated classification of such concepts requires deducing whether their temporal constraints subsume each other, i.e., whether one constraint “matches” the temporal pattern of another because it logically implies it [11]. For example, the concept “infection 1-7 days after surgery” is subsumed by and should be classified below the concept “infection after surgery” because the temporal pattern of the former necessarily implies that of the latter.

Previous researchers in medical informatics have explored the computable representation of abstract temporal relationships between events (i.e., temporal patterns) and the use of such representations in the evaluation or retrieval of patient data. Kohane’s model for temporal reasoning in expert systems defined the logical predicate “Range Relation” (RREL) to represent temporal relationships between medical events and/or states [12]. Although modeled as logical predicates, Range Relations essentially represented linear inequalities between the beginning and/or ending points of time intervals. The syntax of Range Relations allowed the representation of various metric relationships between such intervals, and an inference engine performed pattern matching between Range Relations and stored patient data to retrieve temporally matching instances. The model, however, did not formalize the semantics of Range Relations or the inference operations upon them. Hence, the semantics and computational complexity of such pattern matching remained uncertain.

Later, Das [13,14] defined a formal model to standardize the representation and querying of data stored in time-stamped relational databases, with semantics grounded in the set theory of the relational model. Das’s representation of temporal patterns was limited by not explicitly supporting ranges and metric constraints, such as “within two hours before or four hours after”, and by only retrieving data from relational databases. Subsequent to Das’s work, Shahar defined a general-purpose framework for knowledge-based temporal abstraction, which included a model for representing temporal relationships between intervals and an inference mechanism to retrieve patient data that matched specified patterns [15,16] Shahar’s representation of temporal patterns, like Kohane’s, consisted of linear inequalities over the starting and/or ending points of intervals. But like Kohane, Shahar also did not formalize the semantics of these representations, nor describe the algorithm(s) he implemented to match patient data against such patterns. In the absence of such details, the range of semantics that Shahar’s temporal-matching model could express and the computational complexity of its inferences also remained uncertain. A number of later researchers adopted Shahar’s temporal pattern-matching framework to develop derivative temporal reasoning systems for a variety of specific tasks [17,18], but they, too, did not further formalize or characterize the pattern-matching model. Further, their pattern-matching algorithms were often implemented in opaque software modules that were custom coded for each temporal pattern that needed to be specified [18], which complicated the programming and maintenance of these systems.

Other researchers explored the use of linear inequalities to annotate, in a computable form, the temporal relationships appearing in narrative, unstructured medical documents (such as discharge summaries) [19] and to computably annotate temporal relationships specified in clinical research eligibility criteria [6]. These efforts were valuable in demonstrating that many colloquial concepts of time that appear in narrative data and eligibility criteria can be represented mathematically as linear inequalities, but they did not precisely formalize the semantics of these representations, nor characterize the algorithms needed to correctly match them to each other.

To summarize, the disadvantages of these prior temporal models in the medical domain are that they did not formalize their semantics for representing temporal relationships between pair of events, they did not generalize their temporal pattern-matching algorithms to a single implementation that could handle any represented patterns, and they did not formally prove the soundness or explore the computational complexity of their temporal pattern-matching algorithms.

In this paper, we aim to fill these gaps by formally defining and proving the soundness of (1) a model based on linear inequalities for representing a variety of temporal relationships between pairs of intervals, and (2) an efficient and general-purpose algorithm to deduce whether one temporal relationship represented using this model satisfies (i.e., matches the pattern of, is subsumed by) another temporal relationship.

The described model and algorithm are applicable to a variety of automated temporal pattern-matching and temporal subsumption-testing tasks, including the examples below:

1. Enabling a decision-support system to correctly determine that a specific patient matches the antecedent of the decision-support rule “If infection occurs within two weeks of surgery, …” based on the following structured observation in the patient’s medical record: “The skin infection on the lower abdomen began 10 days after the patient underwent an appendectomy, and resolved two weeks later.”
2. Enabling a data-query application to find all patients at a hospital who were started on mechanical ventilation at some point during an inpatient surgery and remained on ventilation continuously for at least 18 hours after the conclusion of surgery, assuming that the hospital’s database records the start and stop times of all surgical procedures and the start and stop times of all treatments with mechanical ventilation.
3. Enabling medical terminology and ontology systems based on description logic (such as SNOMED-CT [20]) to correctly classify concepts such as the following with respect to each other: “Atrial Fibrillation occurring within 24 hours of surgery”, “Arrhythmia occurring during surgery or within two days after surgery”, “Atrial Fibrillation occurring after surgery”, “Asystole occurring during thoracic surgery”, and “Heart abnormality occurring during surgery or within 1 day after surgery”.

## 2 Materials and Methods

In this section, we describe the model we have defined for representing temporal relationships between pairs of events and the algorithm we have developed to perform temporal pattern matching and temporal subsumption testing with respect to this model.

### 2.1 Examples

Before formally defining the model and algorithm, we present several examples of its use in practice so as to introduce the reader to its syntax and application. In general, the model requires the encoding of temporal relationships between pairs of events so as to represent the semantics of the relationships in a structured, logic-based manner. These encodings then support automated subsumption testing, i.e., the logical deduction of whether one relationship necessarily satisfies (i.e., is subsumed by) another relationship. The examples below are a brief introduction to the model, which is fully explained and formally defined in the sections that follow.

Note that each of the examples entails the representation of temporal relationships between two events, Event_X_ and Event_Y_. Each of the events is assumed to occur over a temporal interval, I_X_ and I_Y_, and each of these intervals is assumed to be defined by a pair of beginning and ending timestamps, b_X_, e_X_ and b_Y_, e_Y_, respectively. For example,

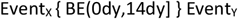

denotes that Event_X_ occurs within 14 days after Event_Y_ (again, the details of the notation are explained in the sections that follow). Each expression between the curly braces in the encodings represents an allowed duration between a pair of timestamps, with the first letter of the expression prefix corresponding to the beginning or ending timestamp of the interval for Event_X_ and the second letter corresponding of the beginning or ending time stamp of the temporal interval for Event_Y_. For example, the expression above denotes that the beginning timestamp of Event_X_ (i.e., b_X_) must be between 0 days and 14 days after the ending timestamp of the Event_Y_ (i.e., e_Y_). In other words, it encodes the temporal constraint: 0 days < b_X_ – e_Y_ <= 14 days. Similarly, the expression

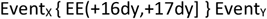

denotes that the ending timestamp of Event_X_ (i.e., e_X_) occurs on the 17^th^ day after the ending timestamp of Event_Y_ (i.e., e_Y_). In other words, it encodes the temporal constraint: 16 days < e_X_ – e_Y_ <= 17 days (or, more colloquially, the temporal relationship “ends 17 days after”).

#### Example 1: Temporal pattern matching of structured narrative data

<u>Temporal Relationship 1</u>:

**“Event**_**X**_ **occurs within two weeks after Event**_**Y**_**”**

<u>Encoding of Temporal Relationship 1</u>:

**Event**_**X**_ **{ BE(0dy**,**14dy] } Event**_**Y**_

<u>Temporal Relationship 2</u>:

**“Event**_**X**_ **started exactly 10 days after Event**_**Y**_, **and ended 1 week later”**

<u>Encoding of Temporal Relationship 2</u>:

**Event**_**X**_ **{ BE[10dy**,**10dy], EE(16dy**,**17dy] } Event**_**Y**_

Based only on these encoded temporal relationships, the algorithm defined in this paper will accurately deduce that Temporal Relationship 2 is subsumed by (i.e., matches the pattern of) Temporal Relationship 1.

#### Example 2: Temporal pattern matching of time-stamped data

<u>Temporal Relationship 1</u>:

**“Event**_**X**_ **occurs within two weeks after Event**_**Y**_**”**

Encoding of Temporal Relationship 1X:

**Event**_**X**_ **{ BE(0dy**,**14dy] } Event**_**Y**_

<u>Temporal Relationship 2</u>:

**“Event**_**X**_ **began on Jan. 16**^**th**^, **2023 at 8:00 AM and Event**_**Y**_ **occurred on Jan. 10**^**th**^, **2023 between 7:30**

**AM and 11:15 AM”**

<u>Encoding of Temporal Relationship 2</u>:

**Event**_**X**_ **{ BE[5.85dy**,**5.85dy] } Event**_**Y**_

Again, based on these encoded temporal relationships, the algorithm defined here will accurately deduce that Temporal Relationship 2 is subsumed by (i.e., matches the pattern of) Temporal Relationship 1. Note that, in this case, the ending timepoint of Event_X_ need not be known to encode and compute subsumption between the relevant temporal relationships.

#### Example 3: Classification and subsumption of temporal concepts in a biomedical terminology/ontology

<u>Concept 1</u>:

**“Pre-surgical antibiotic prophylaxis (within 2 hours prior to surgery)”**

Logical encoding of Concept 1, including temporal relationship:

**Antibiotic Administration { EB[-2hr**,**0hr) } Surgery**

<u>Concept 2</u>:

**“Ceftriaxone antibiotic prophylaxis prior to cholecystectomy (within one hour of procedure)”**

<u>Logical encoding of Concept 2, including temporal relationship</u>:

**Ceftriaxone Administration { EB[-1hr**,**0hr) } Cholecystectomy Surgery**

In this case, the algorithm will again deduce that the temporal relationship encoded in the definition of Concept 1 subsumes the temporal relationship encoded in the definition of Concept 2. Note that, if the concept Antibiotic Administration also subsumes the concept Ceftriaxone Administration per the axioms and inference rules of the terminology/ontology and the concept Surgery likewise subsumes the concept Cholecystectomy Surgery, then Concept 1, in its entirety, subsumes Concept 2 per the rules of logical subsumption in most description logics, such as EL++ [21], which is used to represent the SNOMED-CT terminology and other medical ontologies.

The following sections define the formal syntax and semantics of the temporal model and the subsumption-testing algorithm, and provide additional examples.

### 2.2 Preliminary Assumptions and Definitions

We define a *temporal concept* as a class of events/conditions/states/etc., and an *event* as an instance of a temporal concept. For example, the concept Open-Heart Surgery is a temporal concept, and a specific open-heart surgery procedure performed on a specific patient at a specific time is an event. Note that, in the medical domain, *events* may be symptoms, diseases, procedures, clinical states, or any other phenomena that exist over a defined period of time (i.e., not necessarily “events,” *per se*). For example, the abdominal pain that the patient Mary L. Smith experienced between 7:00 PM on January 21^st^, 2023 and 8:00 AM on January 22^nd^, 2023 is an event that is an instance of the temporal concept Abdominal-Pain.

Each event exists during a finite interval of time, *I* (its *event interval*). Each event interval *I*_X_ is defined by a *beginning timestamp* b_X_ and an *ending timestamp* e_*X*_, where b_X_ and e_X_ are discrete time points on a single fully-ordered linear timeline and -∞ < *b*_*X*_ ≤ *e*_*X*_ < +∞. In the model defined here, these timestamps are specified at a granularity of one second, although variations of the model could specify timestamps at smaller temporal granularities (e.g., one microsecond). The functions b() and e() exist over all temporal intervals *I* and return the beginning and ending timestamps of the intervals, such that b(*I*_*X*_) = b_*X*_ and e(*I*_*X*_) = e_X_. The duration of any interval *I*_*X*_ is equal to e(*I*_*X*_) - b(*I*_*X*_) (or, alternatively, *e*_*X*_ – *b*_*X*_). Note that “instantaneous” events may exist such that b(*I*_*X*_) = e(*I*_*X*_), i.e., the duration of the event = 0.

A *relative temporal duration* is the duration of time between two timestamps, t_1_ and t_2_, which could pertain to two different intervals. This duration may be expressed using any unit of measure greater or equal to the minimum time-point granularity. For example, the following expression denotes the relative temporal duration between the ending timestamp of the temporal interval *I*_X_ and the beginning timestamp of the temporal interval *I*_Y_: e(*I*_X_) – b(*I*_Y_) = -12 hours (“*I*_X_ ended exactly 12 hours before *I*_Y_ began”). Note that relative temporal durations may be positive or negative.

When the minimum time-point granularity is one second, as specified in our model, the allowed units of measure for relative temporal durations are seconds, minutes, hours, days, weeks, months, and years.

Note that the actual beginning and ending timestamps of events need not be known for the model to express temporal relationships between temporal concepts and for the algorithm to make inferences regarding such relationships, specifically subsumption testing. The conceptual existence of temporal intervals with beginning/ending timestamps simply form the formal basis on which the representational model and inference algorithm, including their formal semantics, are based.

### 2.3 Relative Temporal Interval Predicates

A *Relative Temporal Interval Predicate* (RTIP) is a formal predicate expression that denotes a *constraint* on the relative temporal duration between a pair of beginning and ending timestamps belonging to two different temporal intervals. For example, an RTIP might specify that the ending timestamp of the temporal interval corresponding to Event_X_ must precede the beginning timestamp of the temporal interval corresponding to Event_Y_ by a duration greater than 0 days, thereby expressing the constraint “Event_X_ ends before Event_Y_ begins.” We define the syntax and semantics of RTIPs in the following two sections.

### 2.4 RTIP Syntax

RTIP ::= (“BB” | “BE” | “EB” | “EE”) LOWER-BOUND “,” UPPER-BOUND

LOWER-BOUND ::= (FINITE-LOWER-BOUND | INFINITE-LOWER-BOUND)

UPPER-BOUND ::= (FINITE-UPPER-BOUND | INFINITE-UPPER-BOUND)

FINITE-LOWER-BOUND ::= (“[“| “(“) REAL-NUMBER UNIT-OF-MEASURE

INFINITE-LOWER-BOUND ::= “(-INF”

FINITE-UPPER-BOUND ::= REAL-NUMBER UNIT-OF-MEASURE (“)” | “]”)

INFINITE-UPPER-BOUND ::= “+INF)”

REAL-NUMBER ::= Any real number

UNIT-OF-MEASURE ::= (“sc” | “mn” | “hr” | “dy” | “wk” | “mo” | “yr”)

Below are examples of several RTIP expressions and their informal meanings. The formal semantics of RTIP expressions, in general, are presented in the following section.

BB[0dy,+INF) (Event_X_ begins any time after Event_Y_ begins)

BE[1wk,3.5wk] (Event_X_ begins between 1 week and 3.5 weeks after Event_Y_ ends)

EB(-INF,0dy) (Event_X_ ends any time before Event_Y_ begins)

EE[-1dy,1dy] (Event_X_ ends between 1 day before and 1 day after Event_Y_ ends)

Note that *implicit* in the syntactic expression of each RTIP is the pair of temporal intervals over which the predicate is defined, *I*_*X*_ and *I*_*Y*_. For example, the RTIP “BB[0dy,+INF)” actually expresses the predicate “BB[0dy,+INF)[*I*_*X*_,*I*_*Y*_]”, such that, for any pair of temporal intervals *I*_*X*_ and *I*_*Y*_, the RTIP evaluates to either logical TRUE or FALSE.

### 2.5 RTIP Semantics

As mentioned, RTIPs express a constraint on the duration between the beginning or ending timestamps of one temporal interval *I*_X_ and the beginning or ending timestamps of another temporal interval *I*_Y_. The two capitalized letters at the beginning of each RTIP express the pair of timestamps to which the RTIP constraint applies. For any two intervals, there are four combinations of such timestamps, and hence four *types* of RTIPs may be signified by the two capitalized letters. The semantics of the four types are defined as follows:

| RTIP Type | Semantics |
| --- | --- |
| BB | $b(I_x) - b(I_y)$ i.e., Duration between beginning of $I_x$ and beginning of $I_y$ |
| BE | $b(I_x) - e(I_y)$ i.e., Duration between beginning of $I_x$ and ending of $I_y$ |
| EB | $e(I_x) - b(I_y)$ i.e., Duration between ending of $I_x$ and beginning of $I_y$ |
| EE | $e(I_x) - e(I_y)$ i.e., Duration between ending of $I_x$ and ending of $I_y$ |

Figure 1 illustrates the relative temporal durations constrained by each type of RTIP, using two example intervals (recall that relative temporal durations may be negative).

**Figure 1.**
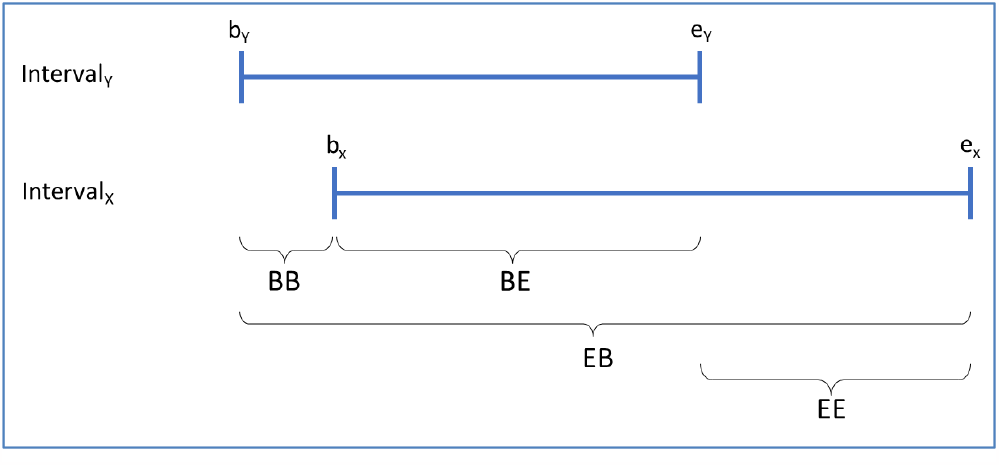
Graphical representation of the four types of RTIPs.

Each RTIP constrains the relative temporal duration between two timestamps to a range of values, and the bounds of this range may assume any finite values between -∞ and +∞ (as well as infinite values, as described below). Finite values of relative temporal duration may be expressed using any of the following temporal units of measure (with their corresponding syntactic tokens): Seconds (“sc”), minutes (“mn”), hours (“hr”), days (“dy”), weeks (“wk”), months (“mn”), and years (“yr”). The model described here establishes the following fixed and consistent conversion factors among these units of measure:

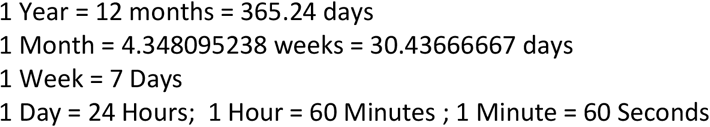

The range specified in each RTIP is defined by a lower and an upper bound, which may each be open or closed. The prefixes “[“and “(“within the lower bound and the suffixes “)” and “]” within the upper bound denote whether the lower and upper bounds of a range are open or closed. Per convention, open bounds are denoted with the characters “(“and “)” and closed bounds with the characters “[“and “]”. The role of these characters in defining the full semantics of RTIPs is illustrated in the following examples (note that the semantics of the special values “(-INF” and “+INF)” will be defined in the following section).

More formally, the range specified by RTIPs of each type has a lower bound L and an upper bound U, such that we can rewrite an RTIP as *ts*_*1*_*ts*_*2*_ ^L, U^ where *ts*_*1*_, *ts*_*2*_ ∈ {B,E} and L,U are valid lower and upper bound expressions, such as “[1wk”, “(-INF”, “1dy]”, “+INF)”, etc. Per this alternative notation, for example, the following RTIP expressions are equivalent: “BB[5dy,+INF)” and “BB^≥ 5dy,< +INF^”. We also define f_*ts*_ as one of the functions “b()” or “e()” that return the beginning or ending timestamp of a temporal interval, respectively (as defined in Section 2.2) (i.e., f_*B*_ = b() and f_*E*_ = e()). We also define the function *value()* to return the value of a lower or upper bound expression, such that, for example, value(“[1wk”) = 1 week and value(“+INF)”) = +∞. In addition, we define the comparison operator “<*” as either one of the mathematical operators “<“or “≤”, depending on whether the corresponding lower or upper bound is open or closed. Lastly, we recall that implicit in the syntactic expression of each RTIP is the pair of temporal intervals, *I*_*X*_ and *I*_*Y*_, over which the predicate is defined.

Using this notation, the formal semantics of an RTIP are defined as follows:

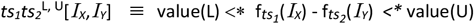

Hence, the formal semantics of

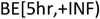

are

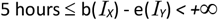

Table 1 contains several examples showing how this general definition indicates the semantics of other specific RTIP expressions.

**Table 1.** Examples of RTIP expressions and their formal semantics.

| RTIP Expression | Semantics |
| --- | --- |
| BB[0dy,+INF) | $0 \text{ days} \leq b(I_X) - b(I_Y) < +\infty$ |
| BE[1wk,3.5wk] | $1 \text{ week} \leq b(I_X) - e(I_Y) \leq 3.5 \text{ weeks}$ |
| EB(-INF,0dy) | $-\infty < e(I_X) - b(I_Y) < 0 \text{ days}$ |
| EE[-1dy,1dy] | $-1 \text{ day} \leq e(I_X) - e(I_Y) \leq 1 \text{ day}$ |

To later assist in defining subsumption over RTIPs, we also define the predicate Satisfies-Bound() to denote that a particular relative temporal duration (*rtd)*, such as e(*I*_*X*_) - b(*I*_*Y*_), satisfies the constraint imposed by a particular lower or upper bound (*L* or *U*) in an RTIP. For example, if an *rtd* = 3 days and *L* = “[1dy”, then Satisfies-Bound*(rtd, L) = TRUE* because 1 day ≤ 3 days. Conversely, if *U* = “-2dy)”, then Satisfies-Bound*(rtd, U) = FALSE* because 3 days ≮-2 days. Using these predicates, we can restate the semantics of an RTIP as follows:

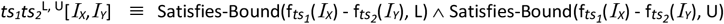

In other words, an RTIP evaluates to TRUE if and only if the indicated relative temporal duration between two intervals satisfies both the lower-bound constraint and the upper-bound constraint of the RTIP. For example,

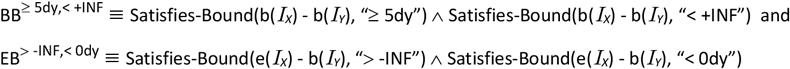

#### 2.5.1 Semantics of “(-INF” and *“*+INF)” in RTIPs

When the special value “(-INF” is used as the lower bound of the range within an RTIP, it means that there is no lower bound on the range, i.e., the specified first timestamp may occur an unlimited duration of time prior to the specified second timestamp. For example, EB(-INF,0dy) denotes that e(*I*_*X*_) - b(*I*_*Y*_) may be an arbitrarily large negative number, i.e., the interval *I*_*X*_ may end any time at all before the interval *I*_*X*_ begins. Conversely, the special value “+INF)” appearing as the upper bound of the range within an RTIP means that there is no upper bound on the range, i.e., the specified first timestamp may occur an unlimited duration of time after the specified second timestamp. Note that these special values are required for RTIPs to express familiar temporal predicates such as “before” and “after,” among others.

### 2.6 Relative Temporal Relationships

Not all useful temporal relationships between pairs of events can be expressed as a single RTIP. For example, expressing the familiar concept of “during” requires the logical conjunction of two RTIPs: BB[0dy,+INF) AND EE(-INF,0dy] (i.e., *I*_*X*_ begins at or after the beginning of *I*_*Y*_ and ends at or before the ending of *I*_*Y*_). As such, we define more generally a *Relative Temporal Relationship* (RTR) between two temporal intervals *I*_*X*_ and *I*_*Y*_ as a logical formula consisting of the conjunction of one or more RTIPs, each defined over the same pair of temporal intervals. We will later generalize this definition to also allow disjunctions of RTIPs within the logical formulae that specify an RTR, but we initially define and prove a model that allows conjunctions only.

### 2.7 RTR Syntax

The syntax of RTRs is defined as follows (with the syntax of “RTIP” remaining as specified above). RTR ::= RTIP (“&” RTIP)*

Example RTR expressions:

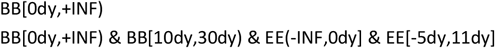

Again, note that *implicit* in the syntactic expression of each constituent RTIP is the pair of temporal intervals over which the predicate is defined, *I*_*X*_ and *I*_*Y*_. For example, the RTR

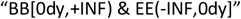

actually expresses the conjoined predicates

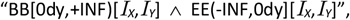

such that, for the single pair of temporal intervals *I*_*X*_ and *I*_*Y*_, the conjunction evaluates to either logical TRUE or FALSE.

Given that all of the RTIPs that specify an RTR are defined over the same pair of temporal intervals, *I*_*X*_ and *I*_*Y*_, it is implicit that the RTR specified by a conjunction of RTIPs is defined over the same pair of intervals, i.e. RTR[*I*_*X*_,*I*_*Y*_]. RTRs are, therefore, themselves predicates that can evaluate to TRUE or FALSE.

### 2.8 RTR Semantics

As noted, an RTR is, itself, a logical predicate defined over a pair of temporal intervals *I*_*X*_ and *I*_*Y*_. The semantics of this predicate are defined as the logical conjunction of the RTR’s constituent RTIPs:

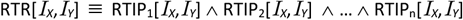

In other words, two temporal intervals satisfy the constraints of an RTR if and only if they satisfy the constraints of all the RTIPs conjoined within the RTR. As a corollary, an RTR can represent any temporal relationship that can be expressed as the conjunction of one or more RTIPs. Although this expressiveness admits a large range of familiar and less-familiar temporal relationships, including “before”, “after”, “during”, “during and after”, and “within 5 hours after,” it cannot express certain other temporal relationships, such as “before or after”. The addition of disjunction to RTR expressions in Section 2.10 will address this limitation, but for the time being we will introduce and formalize the notion of subsumption between RTRs that are limited to conjunctions only. Extending the model to include disjunctions will then follow in a straightforward manner.

### 2.9 Subsumption Between Relative Temporal Relationships

We have thus far defined an RTR as the conjunction of one or more RTIPs, each of which defines a constraint between two temporal intervals, *I*_*X*_ and *I*_*Y*_. We have further defined the semantics of an RTR as the semantics of the logical formula expressed by this conjunction. In other words, two temporal intervals, *I*_*X*_ and *I*_*Y*_, *satisfy* an RTR if they satisfy all of the conjoined predicates that comprise the RTR. For example, two temporal intervals *I*_*X*_ and *I*_*Y*_ satisfy the RTR that denotes “during” (i.e., interval *I*_*X*_ occurs *during* the interval *I*_*Y*_) if they satisfy both the RTIPs “BB[0dy,+INF)” and “EE(-INF,0dy].”

We now define the notion of *subsumption* between two RTRs as follows: RTR_i_ *is subsumed by* RTR_j_ (and RTR_j_ *subsumes by* RTR_i_) if and only if every pair of intervals *I*_*X*_ and *I*_*Y*_ that satisfies the logical formula RTR_i_ necessarily also satisfies RTR_j_. We can denote this relationship as RTR_i_ ⊑ RTR_j_. As an example, we intuitively know that the temporal relationship “during” is subsumed by the temporal relationship “during or after” because any interval *I*_*X*_ that occurs *during* any interval *I*_*Y*_ also necessarily occurs *during or after* the interval *I*_*Y*_. Conversely, we intuitively know that the temporal relationship “before or during” does not subsume the temporal relationship “during or after,” because it is possible that an interval *I*_*X*_ that occurs *before or during* an interval *I*_*Y*_ does not occur *during or after I*_*Y*_ (specifically, if *I*_*X*_ occurs before *I*_*Y*_). Note that, in the latter example, an interval *I*_*X*_ that satisfies “before or during” with respect to *I*_*Y*_ *may* also satisfy “during or after,” but it does not necessarily do so (unlike in the former example, in which satisfaction of “during” by any pair of intervals necessarily implies satisfaction of “during or after”).

The purpose of our work has been to develop a formal symbolic representation model for temporal relationships and an automated inference algorithm that correctly calculates whether any one such temporal relationship subsumes another. As discussed in the introduction, determining whether one temporal relationship between two intervals (or, equivalently, between two events) subsumes another is practically useful for performing temporal pattern matching in the application of decision-support logic or in the analysis of clinical data, as well as for classifying temporally-qualified concepts in biomedical ontologies. Below, we present and prove a logically sound algorithm to test subsumption between any two temporal relationship that are expressed as RTRs (i.e., using the syntax and semantics defined above).

#### 2.9.1 The RTR Subsumption-Testing Algorithm

The algorithm entails the following steps to compute whether a *candidate RTR* (e.g., RTR_i_) is subsumed by a *reference RTR* (e.g., RTR_j_), i.e., whether RTR_i_ ⊑ RTR_j:_

1. Both the candidate and the reference RTRs are *normalized* so that each RTR is converted to a logically equivalent conjunction of exactly four RTIPs, one of each type (i.e., BB, BE, EB, and EE). For example, if RTR_i_ = BB_i_(…) & EE_i_(…) and RTR_j_ = BE_i_(…), the two RTRs are normalized to the following forms (note that the ellipses are placeholders for the constraints specified by each RTIP):

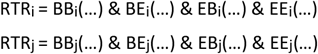

We will address why RTRs are normalized in Section 2.9.2, as well as describe and prove the process to normalize any RTR to a logically equivalent expression of the form above.
2. For each pair of corresponding (same-type) RTIPs in the candidate and reference RTRs, it is determined whether the candidate RTIP is subsumed by the reference RTIP. In other words, with respect to the two normalized RTRs above, the following four subsumption relationships are tested:

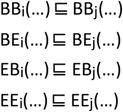

We define subsumption between corresponding RTIPs as follows: RTIP_i_ is subsumed by RTIP_j_ (and RTIP_j_ subsumes RTIP_i_) if and only if (1) RTIP_i_ and RTIP_j_ are of the same type, and (2) all pairs of temporal intervals that satisfy RTIP_i_ also satisfy RTIP_j_. We can formally express this definition for RTIPs of type “BB” (as an example) using logic symbols as follows (the definitions of subsumption for RTIPs of the other three types are analogous):

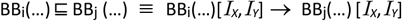

Note that, again, the ellipses in the expressions above are placeholders for the lower and upper bounds of the range specified for each RTIP. The ellipses could be replaced with any valid lower and upper bound expressions, such as in the example below:

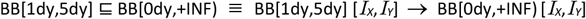

Given this definition of subsumption between RTIPs, subsumption testing with respect to each pair of corresponding (same-type) candidate and reference RTIPs is performed as follows:

Assume the candidate and the reference RTIPs in each corresponding (same-type) pair are represented using the alternative notation for RTIPs introduced in Section 2.5:

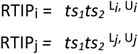

where *ts*_*1*_*ts*_*2*_ ∈ {BB, BE, EB, EE} and L and U are the respective lower and upper bounds of each RTIP.

We can then determine whether the following subsumption is valid

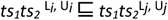

by determining whether the following condition holds:

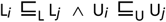

where the comparison operator ⊑_L_ indicates that the lower-bound expression on the lefthand side of the operator is at least as or more restrictive than the lower-bound expression on the righthand side, and the comparison operator ⊑_U_ indicates that the upper-bound expression on the lefthand side of the operator is at least as or more restrictive than the upper-bound on the righthand side. For example, we would test

BB[1dy,5dy] ⊑ BB[0dy,+INF)

by testing whether “[1dy” ⊑_L_ “[0dy” and whether “5dy]” ⊑_U_ “+INF)”

The algorithms to evaluate L_*i*_ ⊑_L_ L_*j*_ and U_*i*_ ⊑_U_ U are defined as follows:

**Evaluating L**_***i***_ **⊑**_L_ **L**_***j***_ :

If L_*i*_ is *-*∞ and L_*j*_ is *-*∞, Then L_*i*_ ⊑_L_ L_*j*_ returns TRUE

Else if L_*i*_ is any finite value and L_*j*_ is *-*∞, Then L_*i*_ ⊑_L_ L_*j*_ returns TRUE

Else if L_*i*_ is *-*∞ and L_*j*_ is any finite value, Then L_*i*_ ⊑_L_ L_*j*_ returns FALSE

Else if both L_*i*_ and L_*j*_ are finite values, Then

If the real number value of L_*i*_ *≥* the real number value of L_*j*_, Then L_*i*_ ⊑_L_ L_*j*_ returns TRUE

Else if the real-number value of L_*i*_ *<* the real-number value of L_*j*_, Then L_*i*_ ⊑_L_ L_*j*_ returns FALSE

Else if the real number value of L_*i*_ = the real number value of L_*j*_, Then

If the comparison symbols of L_*i*_ and L_*j*_ are the same (i.e., both “(“or “[“), Then L_*i*_ ⊑_L_ L_*j*_ returns TRUE

Else if the comparison symbol of L_*i*_ is “(“and that of L_*j*_ is “[“, Then L_*i*_ ⊑_L_ L_*j*_ returns TRUE

Else if the comparison symbol of L_*i*_ is “[“and that of L_*j*_ is “(“, Then L_*i*_ ⊑_L_ L_*j*_ returns FALSE

**Evaluating U**_***i***_ ⊑_U_ **U**_***j***_:

If U_*i*_ is *+*∞ and U_*j*_ is *+*∞, Then U_*i*_ ⊑_U_ U_*j*_ returns TRUE

Else if U_*i*_ is any finite value and U_*j*_ is *+*∞, Then U_*i*_ ⊑_U_ U_*j*_ returns TRUE

Else if U_*i*_ is *+*∞ and U_*j*_ is any finite value, Then U_*i*_ ⊑_U_ U_*j*_ returns FALSE

Else if both U_*i*_ and U_*j*_ are finite values, Then

If the real number value of U_*i*_ ≤ the real number value of U_*j*_, Then U_*i*_ ⊑_U_ U_*j*_ returns TRUE

Else if the real number value of U_*i*_ *>* the real number value of U_*j*_, Then U_*i*_ ⊑_U_ U_*j*_ returns FALSE

Else if the real number value of U_*i*_ = the real number value of U_*j*_, Then

If the comparison symbols of U_*i*_ and U_*j*_ are the same (i.e., both “)” or “]”), Then U_*i*_ ⊑_U_ U_*j*_ returns TRUE

Else if the comparison symbol of U_*i*_ is “)” and that of U_*j*_ is “]”, Then U_*i*_ ⊑_U_ U_*j*_ returns TRUE

Else if the comparison symbol of U_*i*_ is “]” and that of U_*j*_ is “)”, Then U_*i*_ ⊑_U_ U_*j*_ returns FALSE

**Claim 1**: 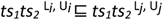 If and only if L_*i*_ ⊑_L_ L_*j*_ ^ U_*i*_ ⊑ U_*j*_, where evaluation of the subsumption predicates “⊑_L_” and “⊑_U_” over pairs of upper and lower bounds, respectively, is performed as defined above.

**Proof of Claim 1:** The details of the proof are provided in the Appendix, Section 7.1. The proof involves the reformulation of Claim 1 using the “Satisfies-Bound()” function defined in Section 2.5 with respect to any relative temporal duration (rtd) and an upper or lower bound, i.e.:

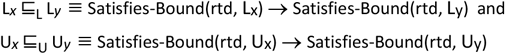

The proof establishes that the evaluation algorithms for the predicates “L_*i*_ ⊑_L_L_*j*_” and “U_*i*_ ⊑_U_ U_*j*_” defined above return TRUE if and only if Satisfies-Bound(rtd, Lx) → Satisfies-Bound(rtd, Ly) and Satisfies-Bound(rtd, Ux) → Satisfies-Bound(rtd, Uy), respectively, for any rtd. For example, the proof establishes that any rtd that satisfies a lower bound consisting of a finite value (such as “> -30 days”) necessarily also satisfies a lower bound consisting of an infinitely negative value (i.e., “> -∞”) because every finite duration (including any finite negative duration) is greater than an infinitely negative duration, per the definition of “infinite.”

3. If all four subsumption relationships between corresponding RTIPs of the candidate and the reference RTRs, RTR_i_ are RTR_j_, are determined to hold (using the testing method described above), the subsumption relationship RTR_i_ ⊑ RTR_j_ is determined to hold.

**Claim 2** (again, using the ellipsis notation for RTIPs):

Assuming

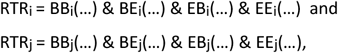

If

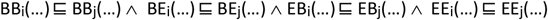

Then

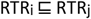

**Proof of Claim 2**: The details of the proof are provided in the Appendix, Section 7.2, but generally depend on the validity of the following propositional logic formula:

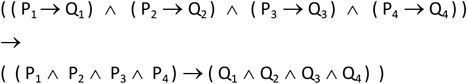

#### 2.9.2 Normalizing RTRs

We have described and proven above how subsumption between any two normalized RTRs can be performed. We now propose that any RTR consisting of the conjunction of one or more RTIPs can be converted to a logically equivalent *normalized RTR* that consists of the conjunction of exactly four RTIPs, one of each type (i.e., BB, BE, EB, and EE). For example, the following RTR (expressed using our alternative notation)

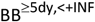

can be converted to the logically equivalent normalized form

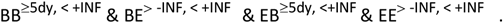

The steps for this conversion process are described and proven below. However, it’s important also to explain the need for such normalization.

Normalization of the candidate RTR and the reference RTR are needed so that subsumption testing between the two RTRs may be performed via the straightforward pair-wise subsumption testing of their corresponding (same-type) RTIPs. In the absence of normalization, certain pairs of candidate/reference RTRs would not have the same set of RTIPs to compare, requiring additional logical inference steps to correctly infer subsumption. For example, consider the two RTRs

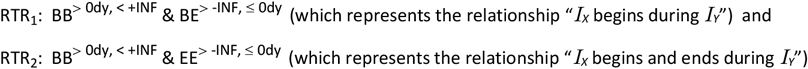

Although it’s intuitively clear that the relationship “*I*_*X*_ begins during *I*_*Y*_” should subsume the relationship “*I*_*X*_ begins and ends during *I*_*Y*_”, the two RTRs are not defined by the same sets of corresponding (matching-type) RTIPs, and, therefore, are not amenable to the complete pair-wise subsumption testing process described in Section 2.9.1. However, because the beginning timestamp of any interval cannot occur later than the ending timestamp of that interval, certain *implications* among RTIPs exist.

Specifically, in the examples above, the following implications hold:

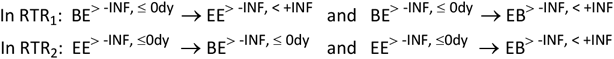

(The enumeration of the all valid implications among RTIPs, in general, appear in Section 2.9.2.1 below.) These implications allow us to rewrite the two RTRs as the logically equivalent expressions:

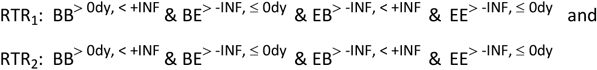

These normalized expressions now do lend themselves to the subsumption testing of RTRs based on the pair-wise subsumption testing of their corresponding (same-type) RTIPs, as was described in Section 2.9.1. Therefore, to optimize subsumption testing, the algorithm first normalizes both the candidate and reference RTRs as an initial step.

In general, the process to normalize an RTR entails the following two sequential operations:

1. Expansion: For each RTIP in the stated definition of the RTR, all of the RTIPs implied by the stated RTIP are generated and conjoined with the stated RTIP. The result is a conjunction of RTIPs that collectively express the equivalent semantics as the original RTR.
2. Consolidation: If the resulting expression includes multiple instances of the same type of RTIP, the conjunction of these RTIPs is replaced by a single RTIP that is logically equivalent to the conjunction. The result is a conjunction of exactly four RTIPs, one of each type (i.e., BB, BE, EB, and EE), which is again semantically equivalent to the original RTR.

##### 2.9.2.1 The Expansion Step in Normalizing RTRs

Table 2 shows the set of valid implications for RTIPs of each type. These implications hold because the ending timestamp of an interval cannot occur before the beginning timestamp, and the beginning timestamp of an interval cannot occur after the ending timestamp.

**Table 2.**
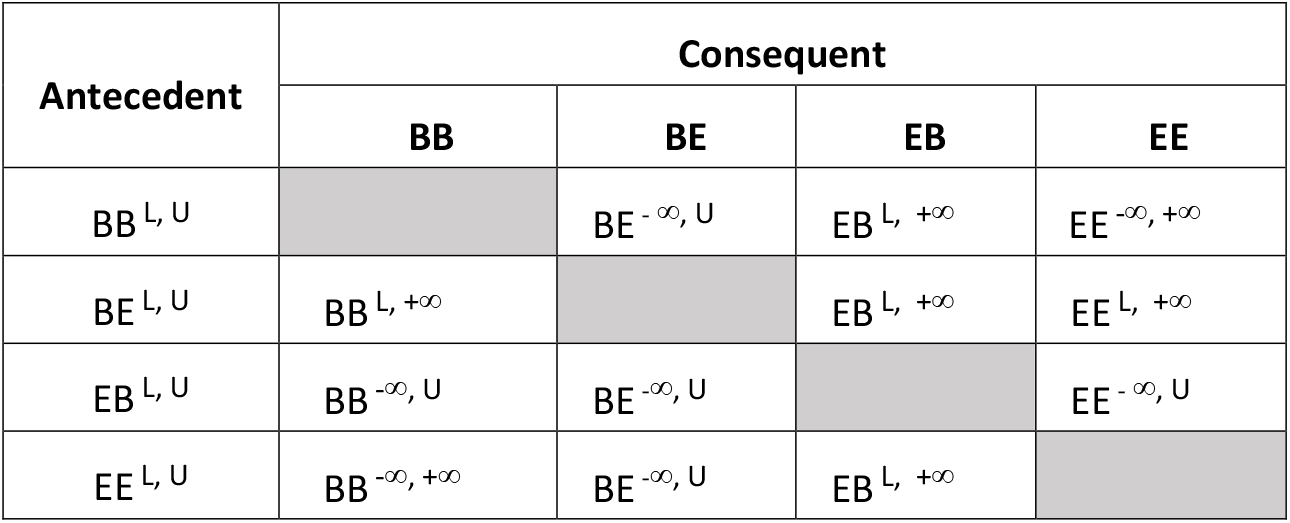
RTIP Implication Table.

| Antecedent | Consequent |  |  |  |
| --- | --- | --- | --- | --- |
|  | BB | BE | EB | EE |
| BB<sup>L, U</sup> |  | BE<sup>-∞, U</sup> | EB<sup>L, +∞</sup> | EE<sup>-∞, +∞</sup> |
| BE<sup>L, U</sup> | BB<sup>L, +∞</sup> |  | EB<sup>L, +∞</sup> | EE<sup>L, +∞</sup> |
| EB<sup>L, U</sup> | BB<sup>-∞, U</sup> | BE<sup>-∞, U</sup> |  | EE<sup>-∞, U</sup> |
| EE<sup>L, U</sup> | BB<sup>-∞, +∞</sup> | BE<sup>-∞, U</sup> | EB<sup>L, +∞</sup> |  |

For example, the following implications regarding specific RTIPs are specified in Table 2:

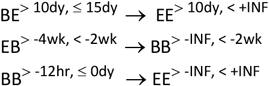

**Claim 3:** Each of the implications specified in Table 2 is valid.

**Proof**: Proofs for the implications in Table 2 appear in the Appendix, Section 0.

The result of expanding a stated RTR based on the implications in Table 2 is another RTR (i.e., another conjunction of RTIPs) that is semantically equivalent to the stated RTR.

**Claim 4**: Any RTR defined by the conjunction of one or more stated RTIPs is logically equivalent to that same conjunction conjoined with any other RTIPs implied by any of the stated RTIPs. I.e.,

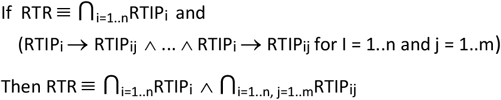

For example, the RTR with a stated definition

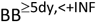

is logically equivalent to

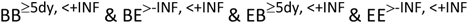

Because of the following implications specified in Table 2:

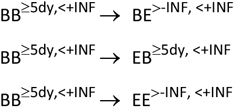

**Proof:** The proof of Claim 4 is provided in the Appendix, Section 7.4.

##### 2.9.2.2 The Consolidation Step in Normalizing RTRs

The expansion step described above may yield a conjunction that includes multiple instances of the same type of RTIP. For example, the following RTR represents the relationship “*I*_*X*_ begins within 5 days of the beginning of *I*_*Y*_ and ends within 2 days of the ending of *I*_*Y*_”

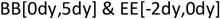

Expanding this expression as described above yields the following conjunction of RTIPs:

BB[0dy,5dy] & EE[-2dy,0dy] & **BE(-INF**,**5dy] & EB[0dy**,**+INF) & EE(-INF**,**+INF) &** <u>BB(-INF,+INF]</u> & <u>BE(-INF,0dy]</u> & <u>EB[-2dy,+INF)</u>

(Note that, for explication only, we have boldfaced the RTIPs implied by the stated RTIP “BB[0dy,5dy]”, and underlined the RTIPs implied by the stated RTIP “EE[-2dy,0dy]”. Refer to Table 1 to confirm these implications).

Rearranging the resulting conjunction, we see that, in this case, it includes two RTIPs of each type:

BB[0dy,+INF) & <u>BB(-INF,+INF]</u> & **BE[-INF**,**+INF) &** <u>BE(-INF,0dy]</u> & **EB[0dy**,**+INF) &** <u>EB(-INF,0dy]</u> & EE(-INF,0dy] & **EE(-INF**,**+INF]**

In general, the expansion of a stated RTR expression may yield arbitrary numbers of RTIPs of the same types, because the definition of an RTR allows for an arbitrary number of stated RTIPs, which each generate three implied RTIPs. As such, to generate the required normalized form for RTRs (i.e., a conjunction including only one RTIP of each type), the subsumption-testing algorithm must consolidate all RTIPs of the same type into a single RTIP that is logically equivalent. The following algorithm accomplishes this consolidation:

For each group (conjunction) of RTIPs of the same type T_x_ (where T_x_ ∈ {BB, BE, EB, EE})

While the conjunction contains more than one RTIP

Calculate the *intersection* of the first two RTIPs in the group, 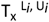 and 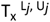, as follows:

Intersection 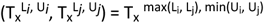

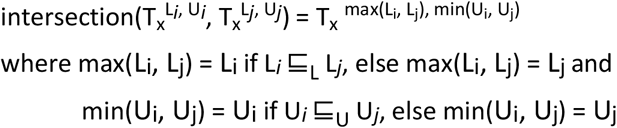

(Note that this calculation uses the same definitions of “⊑_L_” and “⊑_U_” over pairs of lower and upper bounds, respectively, as specified in Section 2.9.1.)

If the resulting intersection is not a valid RTIP, return error “The original RTIP is not valid because no pair of temporal intervals can satisfy it” and quit the subsumption testing algorithm

Replace the first two RTIPs in the group with their intersection

Note that, by the definitions of the subsumption predicates L_*i*_ ⊑_L_ L_*j*_ and U_*i*_ ⊑_U_ U_*j*_ (as provided in Section 2.9.1), the functions max(Li, Lj) and min(Ui, Uj) identify which of the two lower-bound and upper-bound values, respectively, are more restrictive. If a relative temporal duration satisfies the more restrictive lower bound, it necessarily satisfies the less restrictive lower bound, and if a relative temporal duration satisfies the more restrictive upper bound, it necessarily satisfies the less restrictive upper bound.

Therefore, such a relative temporal duration will necessarily satisfy the lower and upper bounds of both of the RTIPs that are being consolidated, and the consolidated RTIP can, therefore, replace the pair of original RTIPs. For example, the intersection of

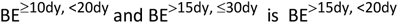

**Claim 5**: For any two RTIPs T_x_^L^^*i*, U*i*^ and T_x_^L^^*j*, U*j*^ where T_x_ ∈ {BB, BE, EB, EE},

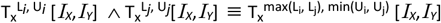

Where max(Li, Lj) = Li if L_*i*_ ⊑_L_ L_*j*_, else max(Li, Lj) = Lj and

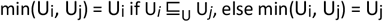

**Proof**: The proof of Claim 5 is provided in the Appendix, Section 7.5. The proof again involves the reformulation of the claim using the “Satisfies-Bound()” function defined in Section 2.5 with respect to any relative temporal duration (rtd) and an upper or lower bound:

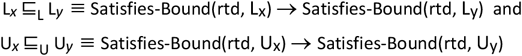

**Claim 6:** Any conjunction of RTIPs of the same type, ⋂_i=1..n_RTIP_i_, can be consolidated to a single RTIP, RTIP_x_, that is logically equivalent to the original conjunction.

**Proof:** If the conjunction consists of only one RTIP, then it is already consolidated. If the conjunction consists of more than one RTIP, then the first two RTIPs can be replaced by their intersection, as calculated above, and the result will be logically equivalent to the prior conjunction, per Claim 4. By induction, this process can be repeated until only a single RTIP remains, which will be logically equivalent to all prior conjunctions, including the original conjunction.

### 2.10 Representing and Reasoning with Temporal Relationships That Include Disjunctions

We have thus far defined a sound algorithm for testing subsumption between temporal relationships that can be represented as a conjunction of RTIPs. We now generalize this algorithm to test subsumption between temporal relationships that can be represented as any combination of conjunctions *and disjunctions* of RTIPs. This generalization is necessary to accommodate common temporal relationships such as “before or after”, as well as more complex relationships, such as “During the first trimester or the third trimester” and “Present at the beginning or the end of”.

We first generalize the syntax of RTRs by adding the disjunction operator, as well as parentheses symbols to designate the intended precedence when conjunctions and disjunctions are combined. The syntax for generalized RTRs (“GRTRs”) is therefore:

GRTR ::= CONJUNCTION-OR-DISJUNCTION ([“&” | “|”] CONJUNCTION-OR-DISJUNCTION)*

CONJUNCTION-OR-DISJUNCTION ::= CONJUNCTION | DISJUNCTION

CONJUNCTION ::= “(“RTIP (“&” RTIP)* “)”

DISJUNCTION ::= “(“RTIP (“|” RTIP)* “)”

Examples of valid GRTR expressions (and their intended meanings):

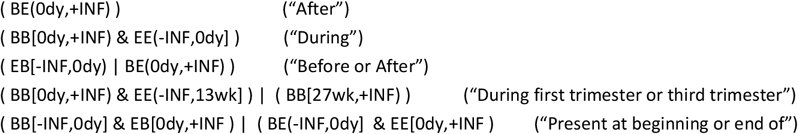

The semantics of RTRs are likewise generalized to reflect that they may now be combinations of disjunctions and conjunctions of RTIPs. To formalize the semantics of such generalized RTRs (GRTRs), first we note that any logical formula consisting of conjunctions and disjunctions of predicates can be converted to a logically equivalent Normal Disjunctive Form (NDF), i.e., a disjunction of conjoined predicates [22]. When all such predicates are RTIPs over pair of the temporal intervals *I*_*X*_ and *I*_*Y*_, we can rewrite any GRTR that has been converted to an NDF as:

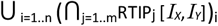

The semantics of a GRTR are, therefore, defined as the NDF of the combination of disjunctions and conjunctions of RTIPs that specifies the GRTR:

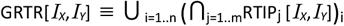

Finally, we generalize the definition of subsumption between any pair of GRTRs as follows: GRTR_a_ is *subsumed by* GRTR_b_ (and GRTR_b_ *subsumes G*RTR_a_) if and only if every pair of intervals *I*_*X*_ and *I*_*Y*_ that satisfies the logical formula GRTR_a_ necessarily also satisfies GRTR_b_. If we again use the symbol ⊑ to denote subsumption, then we define

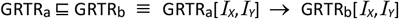

Given the semantics of GRTRs as defined above, we can restate the semantics of subsumption between any pair of GRTRs, GRTR_a_ and GRTR_b_ as follows:

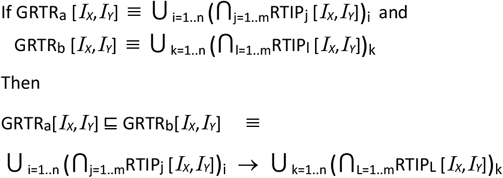

Hence, to test subsumption between GRTRs, we update the subsumption-testing algorithm specified above as follows.

As a first step, convert each GRTR to its Normal Disjunctive Form. Following this conversion, each GRTR will be of the form

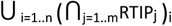

Note that each conjunction in this expression, (⋂_j=1..m_RTIP_j_)_i_, is itself an RTR as defined above (i.e., a conjunction of RTIPs). We can therefore rewrite each GRTR as

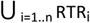

And we can rewrite the definition of subsumption between two GRTRs as GRTR_a_ ⊑ GRTR_b_ ≡

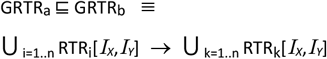

In other words, GRTR_a_ ⊑ GRTR_b_ if and only if the disjunction of RTRs that define GRTR_a_ implies the disjunction of RTRs that implies GRTR_b_. To test whether the implication in the definition above is true, the subsumption algorithm determines (using the subsumption-testing algorithm for RTRs as defined and proven above, including the normalization steps) whether every RTR within the disjunction on the lefthand side of the implication is subsumed by at least one RTR on the righthand side of the implication. In other words, the subsumption algorithm evaluates the truth of the following predicate logic expression:

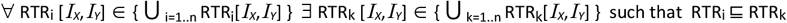

GRTR_a_ **⊑** GRTR_b_ if and only if this logical sentence is TRUE given the definitions of each RTR.

**Claim 7**: For any two GTRTs, GRTR_a_ and GRTR_b_, where

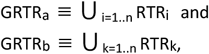

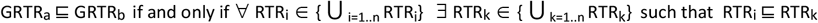

**Proof:** The proof for Claim 7 is provided in the Appendix, Section 7.6

### 2.11 Computational Complexity of the Subsumption-Testing Algorithm

We assert that the computational complexity of testing subsumption between any pair of GRTRs, as described above, is proportional only to the number of disjoined RTRs that appear in the specifications of the two GRTRs. In particular, the number of operations required to test subsumption between two GRTRs in the course of classifying a concept within a terminology/ontology does not at all depend on the size or complexity of the terminology/ontology. Likewise, the number of operations required to test subsumption between two GRTRs in the course of querying an EHR does not at all depend on the number of records in the EHR. Subsumption testing between two GRTRs is a deterministic and “localized” operation, and the number of steps required to perform it depends only on the specification of the two GRTRs, themselves.

Specifically, the number of operations required to perform subsumption testing between two GRTRs is proportional to either (1) the *product* of the number of disjoined RTRs used to specify the candidate and the reference relationships being tested, or (2) the *sum* of the number of disjoined RTRs used to specify the candidate and the reference relationships being tested, whichever is larger.

For example, given the following specifications for two GRTRs:

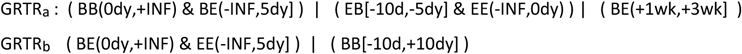

the product of the number of RTRs is 3 x 2 = 6, and the sum of the number of RTRs is 3 + 2 = 5. Hence, the performance of the algorithm to test the proposition GRTR_a_ ⊑ GRTR_b_ is O(6).

In general, if the number of conjoined RTRs in GRTR_a_ is n, and the number of conjoined RTRs in GRTR_b_ is m, then the performance of the operation to test whether GRTR_a_ ⊑ GRTR_b_ is O(max(n*m, n+m)). In the special case that GRTR_a_ and GRTR_b_ each consist of only a single RTR, the performance of the subsumption-testing operation is O(2).

**Proof:** As asserted in Claim 7,

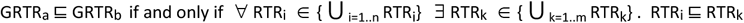

In the worst case, therefore, each RTR_i_ in the specification of the candidate GRTR will need to be subsumption tested against each RTR_k_ in the specification of the reference GRTR to determine whether the quantified logical sentence is true, resulting in no more than n * m subsumption tests of the proposition RTR_i_ ⊑ RTR_k_. Because all RTRs in the specification of both the candidate and the predicate GRTR have been normalized, we know that each test of the proposition RTR_i_ ⊑ RTR_k_ requires exactly four tests of subsumption with respect to the corresponding (same-type) RTIPs in each RTR, i.e., specifically the subsumption tests:

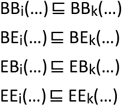

Each such subsumption test of RTIPs is a constant-time operation, as it requires only the comparison of the respective upper and lower bounds of the corresponding candidate and the reference RTIPs, per the algorithms to evaluate ⊑_L_ and ⊑_U_, as specified in Section 2.9.1. Hence, the computational effort of testing whether GRTR_a_ ⊑ GRTR_b_ is proportional to n * m.

In the special cases where (n + m) > (n * m), the computational effort to normalize all of the RTRs in the predicate and candidate GRTRs may be greater than that of testing subsumption between the RTRs in the predicate GRTR and those in the candidate GRTRs. This is because each RTR in the predicate and candidate GRTRs needs to be normalized, and the normalization of each RTR is constant-time in the number of RTIPs that comprise the RTR, which should typically not exceed four (i.e., at most one for each type of RTIP – BB, BE, EB, an EE – as there is no additional information provided in specifying two RTIPs of the same type for an RTR). In the worst case, then, each RTR will consist of four RTIPs, which will each generate (during the expansion step of normalization) three additional RTIPs that are conjoined with those original RTIPs. At most, then, the expansion step will result in a total of 16 RTIPs for each RTR, with a maximum of four RTIPs of each type. Subsequently, up to three merge operations will be needed for each type of RTIP to reduce the four RTIPs to one RTIP during the consolidation step of normalization. Hence, normalizing each RTR will require no more than a total of:

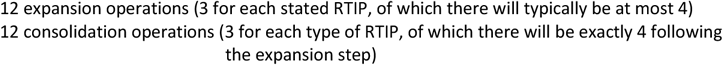

Hence, when (n + m) > (n * m), the total computational effort to test subsumption between two GRTRs may be dominated by the effort required to normalize all of the RTRs within the GRTRs. Note, however, that this situation will only arise when both n and m ≤ 2. However, these cases may be common, so the overall complexity of the subsumption-testing algorithm must be characterized as O(max(n*m, n+m)). In almost all cases, however, both n and m will be < 10, resulting in very efficient performance for the subsumption testing algorithm.

Note, in particular, that the computational complexity of classifying concepts within terminologies/ontologies that use the EL++ description logic (such as SNOMED-CT) is O(n^x^) (i.e., polynomial). Hence, the additional computational effort required to augment the classification process with temporal subsumption (a constant-time operation) will be dominated by the polynomial-time operations already inherent in classification, and therefore will impact the overall performance of EL++ classifiers negligibly.

## 3 Results

This paper describes and formalizes a new, concise, and expressive method for representing temporal relationships between pairs of events and a new, efficient algorithm for testing subsumption between pairs of such relationships. Although the model and algorithm have not yet been implemented and their practical applications have not yet been tested, the proofs provided in this paper substantiate that the subsumption algorithm is sound and correct, based on the defined temporal model. As such, empirical data is not required to substantiate the logical soundness of the model and algorithm.

However, empirical work remains to demonstrate the practical utility of the methods described here, and the paper enumerates several types of practical applications that could benefit from them. These applications include temporal pattern matching (i.e., querying) of semi-structured clinical data that lacks interval timestamps (e.g., data such as “the infection began three days following surgery”) and the classification of temporally-qualified concepts in medical ontologies (e.g., subsumption testing between concepts such as “antibiotic administration prior to surgery” and “ceftriaxone administration within 2 hours prior to cholecystectomy”). The model and algorithm may also be used for temporal pattern matching against time-stamped clinical events, although this task requires first converting timestamped intervals corresponding to pairs of events into relative temporal relationships (RTRs) as defined by our model.

The formalized model and proven algorithm described in this paper form the foundation for applications such as these, among others. We envision incorporation of the algorithm within larger temporal reasoning, data-retrieval, and ontology-management systems as an efficient, modular component specifically for temporal subsumption testing. Future work will investigate these opportunities.

## 4 Discussion

Representing and reasoning with temporal relationships has been a topic of research for a long time, both in the general fields of computer science and logic and in the specific field of medical informatics. A subset of this research has focused on the representation and comparison of temporal intervals.

Allen’s seminal work identified 13 distinct relationships between temporal intervals, including “before”, “during”, “starts”, and “overlaps,” and a set of predicate-logic axioms that govern their behavior (e.g., “before(*I*_*X*_,*I*_*Y*_) & before(*I*_*Y*_,*I*_*Z*_) → before(*I*_*X*_,*I*_*Z*_)”) [1]. Allen argued that these 13 relationships are exhaustive and exclusive, but, in fact, they are not sufficiently expressive to represent important *quantified* relationships between time intervals that exist in the medical domain, such as “Antibiotic administered no more than two hours prior to surgery” or “Infection occurring within 48 after surgery.” The generalized model for representing temporal relationships that we describe can fully represent such quantitative relationships, as well as all 13 of the qualitative relationships defined by Allen. At the same time, the scope of Allen’s overall model for temporal representation and inference is far broader than the work described here, as we have focused solely on representing relationships between pairs of time intervals and computing subsumption between such relationships. There is, perhaps, the potential to augment Allen’s 13 relationships with the more general model for representing temporal relationships described here and to incorporate that enhancement into Allen’s broader model of time and action.

In the medical informatics field, Kohane’s model for temporal reasoning in expert systems defined the logical predicate “Range Relation” (RREL) to represent temporal relationships between medical events and/or states [12]. Like the Relative Temporal Interval Predicates (RTIPs) we have defined, RRELs specified a lower and upper bound for the duration between the beginning and ending points of time intervals corresponding to medical states and/or events. For example, the following predicate in

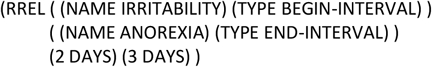

represents the logical assertion that the beginning of irritability precedes the end of anorexia by two to three days. Note that the same assertion would be represented in our model as the following RTIP:

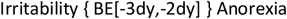

Kohane’s model also allowed for the use of infinitely large as well as infinitesimally small upper and lower bounds to represent qualitative temporal relationships, such as “before” and “after”. For example, the following predicate in his model:

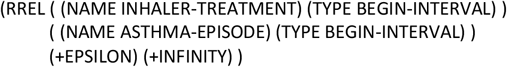

represents the logical assertion that inhaler treatment began after the asthma episode began (he uses “+EPSILON” to denote “a quantity that [the system] takes to be smaller than any numerical value the computer can represent” [12, p. 22]). Note that the same assertion would be represented using our RTIPs as

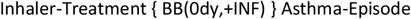

which does not require the introduction of the imprecisely defined value “+EPSILON. Lastly, Kohane’s model also enabled the representation of temporal relationships that require the specification of multiple RRELs, such as

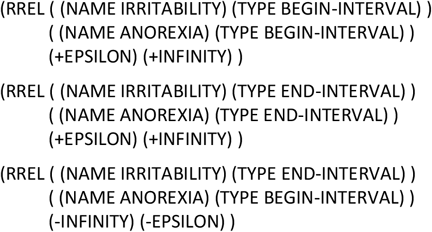

which represents the logical assertion that irritability started prior to but overlapped in time with anorexia. Again, in the model we have described, the same assertion would be represented as a Relative Temporal Relationship (RTR) consisting of a conjunction of three RTIPs:

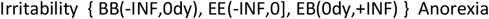

Kohane also defined and implemented a number of inference operations with respect to these representations, including “RREL Retrieval.” This operation performed pattern matching between a specified RREL and instances of stored patient data to retrieve temporally matching instances.

Beyond the greater conciseness of RTRs relative to RRELs, the expressiveness of Kohane’s model for representing temporal relationships among intervals is similar to that of the model we describe. However, unlike in our model, the semantics of RRELs and the operations on them are not formally defined. For example, it is not clear that the three RRELs required to represent the relationship between irritability and anorexia shown above are logically conjoined, and what the implication of conjunction may be for further reasoning and/or the optimization of computations. Further, it is unclear whether

Kohane’s model allows the representation of temporal relationships that require disjunctions of RRELs, as is possible using the model that we describe. Also, the precise semantics of the RREL Retrieval operation and the algorithm(s) to perform it are not defined or described in [12]. The implementation of Kohane’s model (which seems to be based on full first-order predicate logic) may implicitly confer certain semantics and define the extent of temporal expressiveness and inferencing available, but the absence of formal specifications could also lead to unexpected results or intractable computations. In contrast, the constrained and fully defined semantics of the model and algorithm described here formalizes the meaning of all encoded temporal relationships and guarantees near-constant-time performance for subsumption testing between them. At the same time, the overall capabilities of

Kohane’s system for temporal reasoning in medical expert systems goes far beyond those we propose here for the constrained task of subsumption testing between temporal relationships, and he solves numerous challenges that we do not address. Hence, there may, again, be an opportunity to combine both efforts to achieve an incrementally improved system.

Other researchers have addressed temporal representation and reasoning since the work of Kohane. Das [13,14] defined a formal model to standardize the representation and querying of temporal data in relational databases. Like Kohane, Das specified more general inferencing and temporal pattern-matching operations than we do here, as he was concerned with developing an overall “temporal mediator” for intelligent systems. Das, however, did specify formal semantics for his representation and inferencing system, in his case grounded in the set-theoretic foundations of the relational data model.

The inferencing capabilities he specified include temporal pattern matching based on the interval-comparison operations defined by Allen (i.e., “before”, “overlaps”, etc.), although his operators do not explicitly support ranges (unlike ours and Kohane’s) and therefore cannot easily express comparisons such as “within two hours before.” Also unlike our work, Das’s inferencing model is confined to the task of retrieving time-stamped data from relational databases, whereas the model and algorithm we describe allow for pattern matching and subsumption testing with respect to data that are not necessarily timestamped (such as the narrative expression “the infection started four days after the appendectomy”) and between abstract concepts that are not instantiated in a database (for example, pairs of logically defined concepts in a medical terminology or ontology).

Subsequent to Das’s work, Shahar defined a general-purpose framework for knowledge-based temporal abstraction [15,16]. Again, the scope of Shahar’s model far exceeded that of the work described here, although it did include a method for temporal pattern matching to identify temporal relationships in patient data and/or retrieve patient data that matched certain relationships. Like the models defined by Kohane, Das, and ourselves, Shahar’s representation system recognized that temporal relationships such as “during” and “after” can be formally represented as constraints with respect to the beginning and ending time points of temporal intervals. Specifically, he defined a data structure called a “dynamic induction relation of a context interval (DIRC),” which could be used to specify temporal patterns by constraining the four types of relationships between the respective end points of two intervals (i.e., “ss”, “se”, “es”, and “ee,” denoting “starting” and “ending” timepoints), analogous to the four types of relationships that can be expressed by RTIPs in our own model (i.e., BB, BE, EB, and EE). However, beyond this high-level conceptualization, Shahar’s work did not formalize the semantics of DIRCs and the temporal patterns they can represent, nor describe the algorithm(s) he implemented to match patient data against such patterns or compare such patterns to each other (e.g., to test subsumption).

The work described here has formalized the types of temporal relationships that our model can represent, as well as defined and proven an efficient algorithm for performing temporal pattern matching and subsumption testing. Again, given the broader scope and greater overall capabilities of Shahar’s framework for knowledge-based temporal reasoning, it is possible that our work could be incorporated into his as a more completely formalized and optimized “sub-system” for the specific temporal representation and inferencing that our model and algorithm support.

## 5 Conclusion

We have formally defined the syntax and semantics of a model for representing relative temporal relationships between pairs of medical events, such as disorders, symptoms, physical exam findings, procedures, and treatments. Based on this model, we have described a sound, logic-based algorithm to compute subsumption between any pair of such temporal relationships, the use of which can support temporal pattern matching against time-stamped or semi-structured data, as well as classification of concepts in logic-based terminologies and ontologies. Our proofs of the algorithm’s correctness, based on the nature of temporal intervals and the axioms of propositional logic, ensure that applications of the algorithm will yield sound and correct results. We have also shown that the algorithm has constant-time computational complexity, which varies only with the complexity of the temporal relationships expressed, rather than with the size of the ontology or database to which the algorithm is applied. Although related to previous models for representing and computing with temporal relationships (particularly, intervals), our model is more formalized and our algorithm has been formally proven. The scope and effectiveness of our methods, therefore, are well defined. Given the relatively narrow scope of our temporal model and inferencing algorithm, however, we anticipate that the work described here will be best applied as a sub-component of larger systems for data retrieval and knowledge management.

## Data Availability

No data was produced in the present work to date

## Acknowledgements

This work was funded by the Systemic Harmonization and Interoperability Enhancement for Lab Data (SHIELD) Program, Center for Devices and Radiologic Health, U.S. Food and Drug Administration (FDA Contract 2103911).

## 7 Appendix A

### 7.1 Proof of Claim 1: Testing Subsumption Between RTIPs of the Same Type

**Claim 1**: If 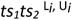 and 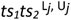 represent two valid RTIPs of the same type (where *ts*_*1*_*ts*_*2*_ ∈ {BB, BE, EB, EE}, then 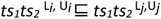 if and only if L_*i*_ ⊑_L_ L_*j*_ ^ U_*i*_ ⊑_U_ U_*j*_, where evaluation of the subsumption predicates “⊑_L_” and “⊑_U_” over pairs of upper and lower bounds, respectively, are performed using the following algorithm:

**Evaluating L**_***i***_ **⊑**_L_ **L**_***j***_ :

If L_*i*_ is *-*∞ and L_*j*_ is *-*∞, Then L_*i*_ ⊑_L_ L_*j*_ returns TRUE

Else if L_*i*_ is any finite value and L_*j*_ is *-*∞, Then L_*i*_ ⊑_L_ L_*j*_ returns TRUE

Else if L_*i*_ is *-*∞ and L_*j*_ is any finite value, Then L_*i*_ ⊑_L_ L_*j*_ returns FALSE

Else if both L_*i*_ and L_*j*_ are finite values, Then

Else if the real number value of L_*i*_ = the real number value of L_*j*_, Then

(There are no other cases)

**Evaluating U**_***i***_ ⊑_U_ **U**_***j***_:

If U_*i*_ is *+*∞ and U_*j*_ is *+*∞, Then U_*i*_ ⊑_U_ U_*j*_ returns TRUE

Else if U_*i*_ is any finite value and U_*j*_ is *+*∞, Then U_*i*_ ⊑_U_ U_*j*_ returns TRUE

Else if U_*i*_ is *+*∞ and U_*j*_ is any finite value, Then U_*i*_ ⊑_U_ U_*j*_ returns FALSE

Else if both U_*i*_ and U_*j*_ are finite values, Then

Else if the real number value of U_*i*_ = the real number value of U_*j*_, Then

If the comparison symbols of U_*i*_ and U_*j*_ are the same (i.e., both “)” or “]”), Then U_*i*_ ⊑_U_ U_*j*_ returns TRUE Else if the comparison symbol of U_*i*_ is “)” and that of U_*j*_ is “]”, Then U_*i*_ ⊑_U_ U_*j*_ returns TRUE

(There are no other cases)

**Proof of Claim 1:**

Recall that, in Section 2.5, we defined the formal semantics of an RTIP as

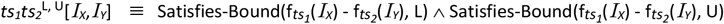

where

1. *ts*_*1*_, *ts*_*2*_ ∈ {B,E}
2. L and U are valid lower-bound and upper-bound expressions, such as “[1wk”, “(-INF”, “1dy]”, “+INF)”, etc.
3. f_*ts*_ is one of the functions “b()” or “e()” that returns the beginning or ending timestamp of a temporal interval *I*_*X*_, such that b(*I*_*X*_) = b_*X*_ and e(*I*_*X*_) = e_X_. The specific function that f_*ts*_ represents depends on whether ts = “B” or “E,” such that f_*B*_ = b() and f_*E*_ = e().
4. The predicate Satisfies-Bound(<rtd>,<L or U>) denotes that a particular relative temporal duration (*rtd)* such as “b_*X*_ – e_Y_” satisfies the constraint imposed by a particular lower or upper bound (*L* or *U*) in an RTIP. For example, Satisfies-Bound(3 days, *L* = “[1 day”) *= TRUE* because 3 days ≥ 1 day.

Also, in Section 2.9.1, we defined the semantics of subsumption between two RTIPs as: RTIP_i_ is subsumed by RTIP_j_ (and RTIP_j_ subsumes RTIP_i_) if and only if (1) RTIP_i_ and RTIP_j_ are of the same type, and (2) all pairs of temporal intervals that satisfy RTIP_i_ also satisfy RTIP_j_. Based on the formal semantics of RTIPs as defined above, this definition of subsumption can be expressed as

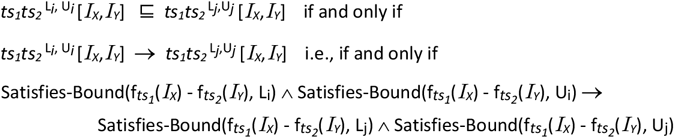

Because the types of both RTIPs must be the same (per the assumptions of Claim 1), we can replace each instance of “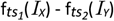” in the expression above with “rrtd[*I*_*X*_,*I*_*Y*_]” to represent the value of the *relevant relative temporal duration* expressed by “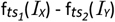” (for example, rrtd[*I*_*X*_,*I*_*Y*_] = b(*I*_*X*_) - e(*I*_*Y*_) if the two RTIPs are both of the type “BE”). As such, the definition of subsumption between two RTIPs can be re-expressed as

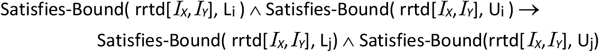

By propositional logic, we know this expression holds if the following expression holds

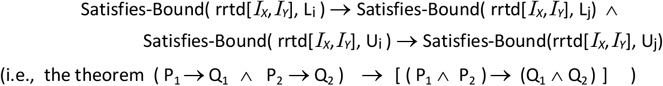

Hence, we only need to show that

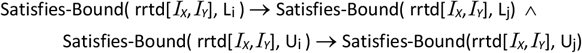

to show that

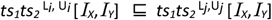

Given this restatement of Claim 1, we only need to prove that the algorithms for evaluating the subsumption predicates L_*i*_ ⊑_L_ L_*j*_ and U_*i*_ ⊑_U_ U_*j*_ are equivalent to evaluating the expressions [Satisfies-Bound(rrtd[*I*_*X*_,*I*_*Y*_], L_i_) → Satisfies-Bound(rrtd[*I*_*X*_,*I*_*Y*_], L_j_)] and [Satisfies-Bound(rrtd[*I*_*X*_,*I*_*Y*_], U_i_) → Satisfies-Bound(rrtd[*I*_*X*_,*I*_*Y*_], U_j_)], respectively, for any rrtd[*I*_*X*_,*I*_*Y*_]. We prove this assertion by proving the validity of each step in the algorithms.

**Evaluating L**_***i***_ **⊑**_L_ **L**_***j***_ :

If L_*i*_ is *-*∞ and L_*j*_ is *-*∞, Then L_*i*_ ⊑_L_ L_*j*_ returns TRUE

*Any rrtd that is > -*∞ *is necessarily > -*∞, *by tautology*

Else if L_*i*_ is any finite value and L_*j*_ is *-*∞, Then L_*i*_ ⊑_L_ L_*j*_ returns TRUE

*Any rrtd that is > or ≥ a finite value is necessarily > -*∞, *because -*∞ *< every finite value, by definition*

Else if L_*i*_ is *-*∞ and L_*j*_ is any finite value, Then L_*i*_ ⊑_L_ L_*j*_ returns FALSE

*Assume that the finite value of* L_*j*_ = X. *Because -*∞ *< every finite value (by definition), there also exists a finite value Y such that -*∞ *< Y < X. If rrtd* =*Y, then rrtd > -*∞ *(satisfying* L_*i*_*) but rrtd < X (not satisfying* L_*j*_, *regardless of whether it is open (“> X “) or closed (“≥ X”))*.

Else if both L_*i*_ and L_*j*_ are finite values, Then

If the real-number of L_*i*_ *≥* the real-number of L_*j*_, Then L_*i*_ ⊑_L_ L_*j*_ returns TRUE

*Assume Y is the finite value of* L_*i*_ *and Z is the finite value of* L_*j*_. *If Y ≥ Z, any rrtd X that is > or ≥ Y is necessarily > and ≥ Z (respectively) by the transitivity properties of > and ≥*.

Else if the real-number value of L_*i*_ *<* the real-number value of L_*j*_, Then L_*i*_ ⊑_L_ L_*j*_ returns FALSE *Assume Y is the real-number value of* L_*i*_ *and Z is the real-number value of* L_*j*_. *If Y < Z, then there exists an rrtd X that is > or ≥ Y (satisfying* L_*i*_*), but < Z (not satisfying* L_*j*_, *regardless of whether Z is open (“> Z”) or closed (“≥ Z”). For example, an rrtd that is 1 second < Z would satisfy* L_*i*_ *but not* L_*j*_.

Else if the real number value of L_*i*_ = the real number value of L_*j*_, Then

*If the real number values and the comparison symbols of* L_*i*_ and L_*j*_ *are both the same, then* L_*i*_ = L_*j*_. *In this case, if any rrtd satisfies* L_*i*_, it necessarily satisfies L_*j*_ by tautology.

*If any rrtd X is > a finite value Y, then it is necessarily ≥ Y (give the logical meaning of “≥” – i*.*e*., *“> OR* = *“, and the propositional logic theorem* P→ (P ∨ Q).

*Assume the finite value of both* L_*i*_ and L_*j*_ = *Y and the value of rrtd* = *X. If X* = *Y, then the rrtd satisfies* L_*i*_, *(“≥ Y”), but does not satisfy* L_*j*_ *(“> Y”)*.

(There are no other cases)

**Evaluating U**_***i***_ ⊑_U_ **U**_***j***_: The proofs of the algorithmic steps for Evaluating U_*i*_ ⊑_U_ U_*j*_ are all converses of the proofs of the steps for Evaluating L_*i*_ ⊑_L_ L_*j*_ presented above, so we do not repeat them here.

### 7.2 Proof of Claim 2: Subsumption Testing Between Normalized RTRs

**Claim 2:**

Assuming

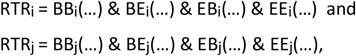

If

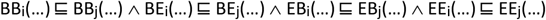

Then

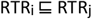

Note that the ellipses in the expressions above are placeholders for the lower and upper bounds of the range specified for each RTIP. The ellipses could be replaced with any valid lower and upper bound expressions, such as in the example below:

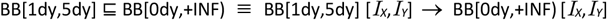

**Proof:**

The proof of Claim 2 follows from the definitions of RTRs and RTIPs and the semantics of subsumption with respect to RTRs and RTIPs. First, based on the definition of an RTR, we can rewrite the normalized definitions of RTR_i_ and RTR_j_ as the following logical definitions:

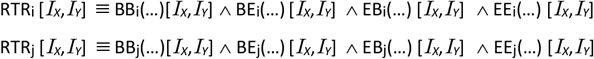

Then, based on these logical equivalences and the semantics of subsumption with respect to RTRs and RTIPs, we can rewrite Claim 2 as the following logical formula

If

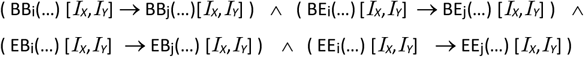

Then

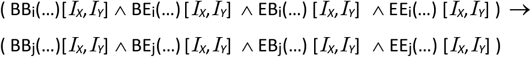

Note that this formula is equivalent to the propositional logic statement

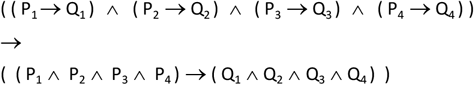

We prove this statement by proving the more general proposition:

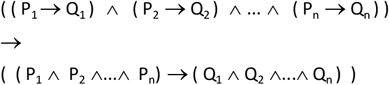

which can be restated as

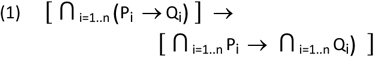

We prove (1) by induction. The base case is (1) with n=1, i.e.

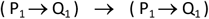

which we can re-write as below in our induction proof.

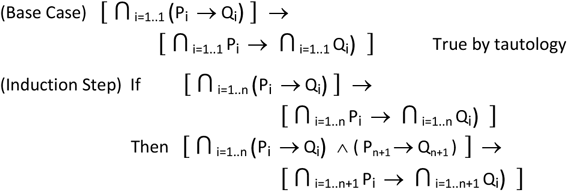

Proof:

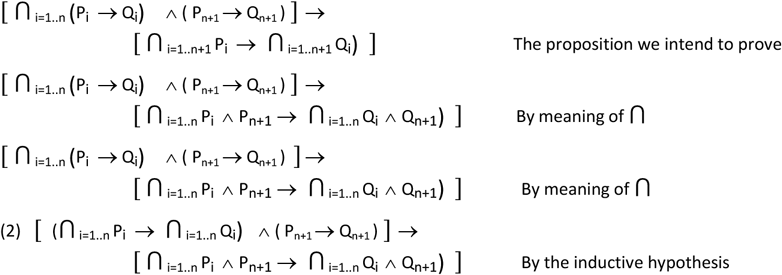

Now we make the following substitutions of proposition symbols in statement (2) above to improve clarity of the remaining proof steps:

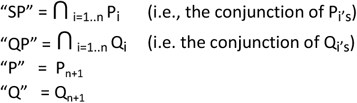

The previous proposition (2) can then be re-written as:

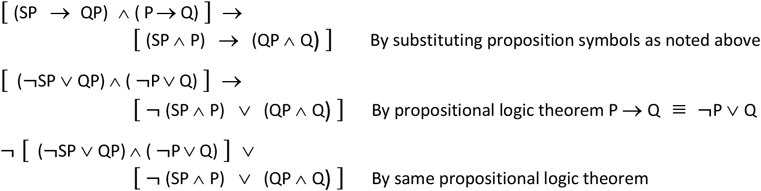

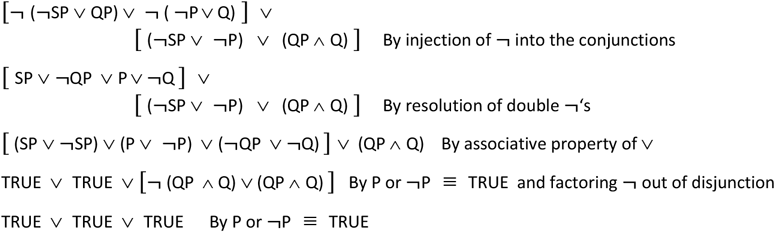

We have therefore proven Claim 2.

### 7.3 Proof of Claim 3: Validity of the Implications Listed in Table 2

The proofs for each cell of the implications table follow from the definition of intervals in Section 2.2, specifically that the *beginning timestamp* b_X_ and the *ending timestamp e*_*X*_ of any event interval *I*_*X*_ are discrete time points on a single fully-ordered linear timeline such that -∞ < *b*_*X*_ ≤ *e*_*X*_ < +∞. This definition implies that ending timestamp of an interval cannot occur before the beginning timestamp, and the beginning timestamp of an interval cannot occur after the ending timestamp. It also implies that the ending timestamp of an interval may occur at any point after the beginning timestamp (up to an infinite duration of time afterwards), and the beginning timestamp of an interval may occur at any point before the ending timestamp (up to an infinite duration of time beforehand). These implications can be formally represented as

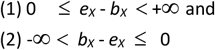

for any interval *I*_*X*_. For any interval *I*_*Y*_, of course, the same implications exist with the subscript “x” replaced by the subscript “y”.

Recall also from Section 2.5 that we can rewrite an RTIP as *ts*_*1*_*ts*_*2*_ ^L, U^ where *ts*_*1*_, *ts*_*2*_ ∈ {B,E} and L,U are valid lower and upper bound expressions, such as “[1wk”, “(-INF”, “1dy]”, “+INF)”, etc. We also defined f_*ts*_ as one of the functions “b()” or “e()” over a temporal interval that return the beginning or ending timestamp of that interval, such that f_*B*_ = b(*I*_*X*_) = *b*_*X*_ and f_*E*_ = e(*I*_*X*_) = *e*_*X*_. We also defined the function value() to return the value of a lower or upper bound expression in an RTIP, such that, for example, value(“[1wk”) = 1 week and value(“+INF)”) = *+*∞. In addition, we defined the comparison operator “<*” as one of the mathematical operators “<“or “≤”, depending on whether the corresponding lower or upper bound, L or U, is open or closed. Lastly, we noted that, implicit in the syntactic expression of each RTIP, is the pair of temporal intervals, *I*_*X*_ and *I*_*Y*_, over which the predicate is defined.

Using these definition and notations, we then defined the formal semantics of an RTIP as follows:

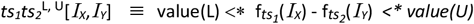

For example,, the formal semantics of BE^L, U^[*I*_*X*_,*I*_*Y*_] ≡ value(L) <* *b*_*X*_ - *e*_*X*_ *<* value(U)*.

Given these definitions and notation, we can derive each of the implications in Table 2 of Section 2.9.2.1 by simply adding the inequalities representing the antecedents in the table to the inequalities (1) and/or listed above, which are implied by our model assumptions. The sum of these inequalities yields the consequents specified in Table 3 below. Each cell in Table 3 is, therefore, an instance a proof in the following form (using the BB^L,U^ → EB ^-∞,U^ cell as an example):

If

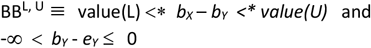

Then

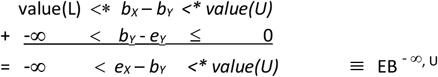

Note that, in the table below, we abbreviate the “value()” function as “v()”.

**Table 3.** Proofs of the implications for RTIPs.

| Ante-<br>cedent | Consequent |  |  |  |
| --- | --- | --- | --- | --- |
|  | BB | BE | EB | EE |
| $BB^{L,U}$ | | $\begin{array}{l} v(L) <^* b_X - b_Y <^* v(U) \\ + \frac{-\infty < b_Y - e_Y \leq 0}{-\infty < b_X - e_Y <^* v(U)} \end{array}$ | $\begin{array}{l} v(L) <^* b_X - b_Y <^* v(U) \\ + \frac{0 \leq e_X - b_X < +\infty}{v(L) <^* e_X - b_Y < +\infty} \end{array}$ | $\begin{array}{l} v(L) <^* b_X - b_Y <^* v(U) \\ -\infty \leq b_Y - e_Y < 0 \\ + \frac{0 \leq e_X - b_X < +\infty}{-\infty < e_X - e_Y < +\infty} \end{array}$ |
| $BE^{L,U}$ | $\begin{array}{l} v(L) <^* b_X - e_Y <^* v(U) \\ + \frac{0 \leq e_Y - b_Y < +\infty}{v(L) <^* b_X - b_Y < +\infty} \end{array}$ | | $\begin{array}{l} v(L) <^* b_X - e_Y <^* v(U) \\ 0 \leq e_Y - b_Y < +\infty \\ + \frac{0 \leq e_X - b_X < +\infty}{v(L) <^* e_X - b_Y < +\infty} \end{array}$ | $\begin{array}{l} v(L) <^* b_X - e_Y <^* v(U) \\ + \frac{0 \leq e_X - b_X < +\infty}{v(L) <^* e_X - e_Y < +\infty} \end{array}$ |
| $EB^{L,U}$ | $\begin{array}{l} v(L) <^* e_X - b_Y <^* v(U) \\ + \frac{-\infty < b_X - e_X \leq 0}{-\infty < b_X - b_Y <^* v(U)} \end{array}$ | $\begin{array}{l} v(L) <^* e_X - b_Y <^* v(U) \\ -\infty < b_X - e_X \leq 0 \\ + \frac{-\infty < b_Y - e_Y \leq 0}{-\infty < b_X - e_Y <^* v(U)} \end{array}$ | | $\begin{array}{l} v(L) <^* e_X - b_Y <^* v(U) \\ + \frac{-\infty < b_Y - e_Y \leq 0}{-\infty < e_X - e_Y <^* v(U)} \end{array}$ |
| $EE^{L,U}$ | $\begin{array}{l} v(L) <^* e_X - e_Y <^* v(U) \\ 0 \leq e_Y - b_Y < +\infty \\ + \frac{-\infty < b_X - e_X \leq 0}{-\infty < b_X - b_Y < +\infty} \end{array}$ | $\begin{array}{l} v(L) <^* e_X - e_Y <^* v(U) \\ + \frac{-\infty < b_X - e_X \leq 0}{-\infty < b_X - e_Y <^* v(U)} \end{array}$ | $\begin{array}{l} v(L) <^* e_X - e_Y <^* v(U) \\ + \frac{0 \leq e_Y - b_Y < +\infty}{v(L) <^* e_X - b_Y < +\infty} \end{array}$ | |

Also, note the following properties for additions that appear in the table (where “RN” is any real number):

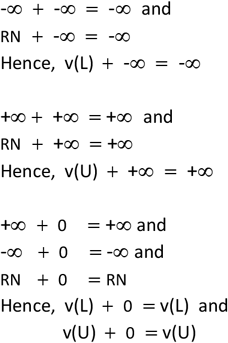

Lastly, note the following properties of inequalities when they are added, which allow us to combine “<“, “≤”, and “<*” as shown in the additions below:

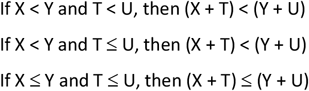

### 7.4 Proof of Claim 4: Equivalence of Stated and Expanded RTRs

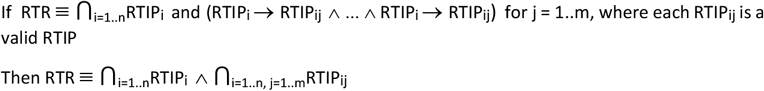

Note that Claim 3 is equivalent to

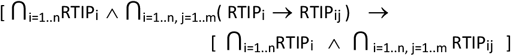

For brevity and clarity, we replace each instance of RTIP_i_ with P_i_ and RTIP_ij_ with R_ij_, so Claim 3 becomes

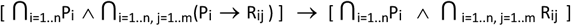

**Proof:** We begin by proving the special case of the Claim when n=1, which can be rewritten as

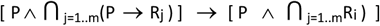

We prove this case by induction, beginning with the base case where j = 1:

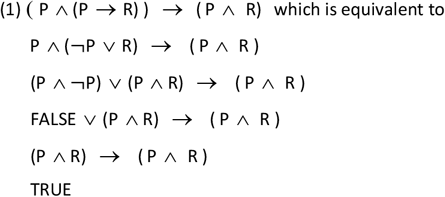

Hence, we have proven (P ^ (P → R)) → (P ^ R), which is equivalent to P ^ ⋂_j=1..1_(P → R_j_) → (P ^ ⋂_j=1..1_R_j_) [the base case in the following inductive proof]

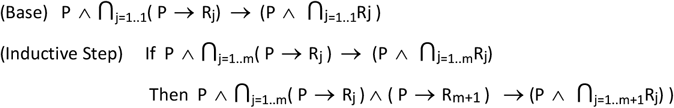

Proof:

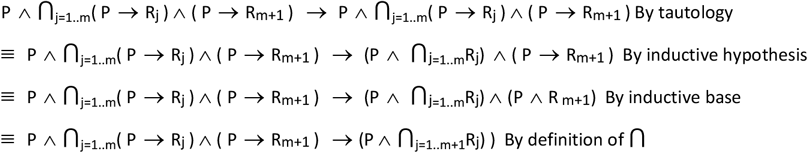

Hence, we have shows that, for each RTIP_i_ in Claim 3,

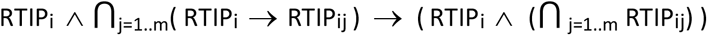

Hence, conjoining over all RTIPs in the RTR,

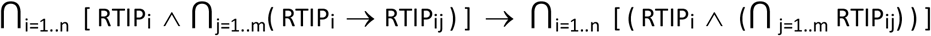

Finally, by the associativity property of conjunction,

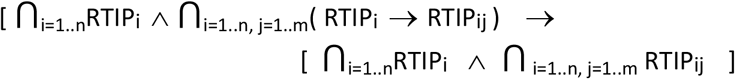

Hence, we have proven Claim 4.

#### 7.5 Proof of Claim 5: Equivalence of the Intersection of Two RTIPs

**Claim 5**: For any two RTIPs T_x_^L^*i*^, U*i*^ and T_x_^L^^*j*, U*j*^ where T_x_ ∈ {BB, BE, EB, EE},

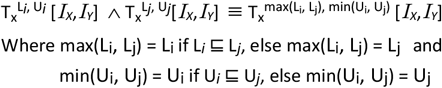

**Proof:** Recall the definition of the semantics of an RTIP:

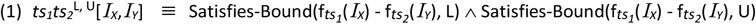

Given a particular type of RTIP we can rewrite the expression “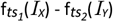” in (1) as “rrtd(*I*_*X*_,*I*_*Y*_)”, which represents the relevant relative temporal duration (rrtd) function with respect to *I*_*X*_,*I*_*Y*_ that is indicated by type of RTIP *ts*_*1*_*ts*_*2*_. For example, if *ts*_*1*_*ts*_*2*_ = “BE”, then rrtd(*I*_*X*_,*I*_*Y*_) = b(*I*_*X*_) - e(*I*_*Y*_), whereas if *ts*_*1*_*ts*_*2*_ = “EE”, then rrtd(*I*_*X*_,*I*_*Y*_) = e(*I*_*X*_) - e(*I*_*Y*_). For brevity, we can also replace *ts*_*1*_*ts*_*2*_ in this definition by T_x_ where T_x_ ∈ {BB, BE, EB, EE}. We can then rewrite (1) as the equivalent definition

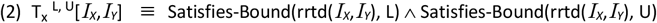

Using (2) and the rules of propositional logic, we can define the conjunction of two RTIPs as:

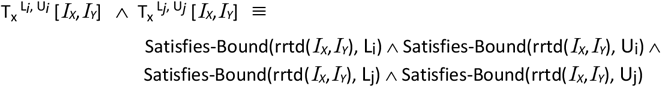

Rearranging the conjunction on the right yields

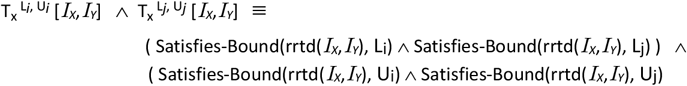

Using (2) we can also assert

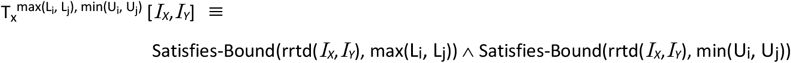

Hence, by transitivity of logical equivalence, proving Claim 4 can be accomplished by proving the following logical formula:

(3) (Satisfies-Bound(rrtd(*I*_*X*_,*I*_*Y*_), Li) ^ Satisfies-Bound(rrtd(*I*_*X*_,*I*_*Y*_), Lj)) ^

(Satisfies-Bound(rrtd(*I*_*X*_,*I*_*Y*_), Ui) ^ Satisfies-Bound(rrtd(*I*_*X*_,*I*_*Y*_), Uj)

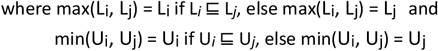

Proving (3) requires proving both of the logical formulae (4) and (5) below:

(4) (Satisfies-Bound(rrtd(*I*_*X*_,*I*_*Y*_), Li) ^ Satisfies-Bound(rrtd(*I*_*X*_,*I*_*Y*_), Lj) ^

Satisfies-Bound(rrtd(*I*_*X*_,*I*_*Y*_), Ui) ^ Satisfies-Bound(rrtd(*I*_*X*_,*I*_*Y*_), Uj))

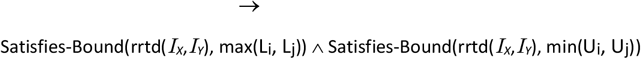

(5) Satisfies-Bound(rrtd(*I*_*X*_,*I*_*Y*_), max(Li, Lj)) ^ Satisfies-Bound(rrtd(*I*_*X*_,*I*_*Y*_), min(Ui, Uj))

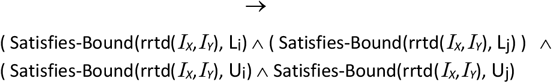

To prove (4), note that max(Li, Lj) must be either Li or Lj. Regardless of which it is (i.e., substituting either Li or Lj for max(Li, Lj)), we know that

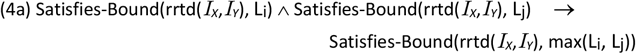

because, in general, the propositional formulae P ^ Q → P and P ^ Q → Q are both TRUE.

The same argument applies to min(Ui, Uj), i.e., it must be either Ui or Uj. Hence, by the same propositional logic, we also know that

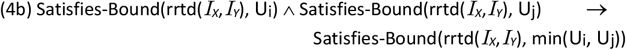

Hence, we can combine (4a) and (4b) to prove (4), because, in general, the following proposition holds:

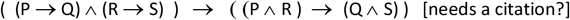

To prove (5), note again that max(Li, Lj) must be either Li or Lj.

If max(Li, Lj) = Li, we know (by the definitions of max()) that Li ⊑ Lj, and therefore (by the definition of the predicate “⊑” and substitution of max(Li, Lj) for Li in that definition), that

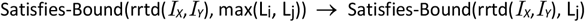

Because max(Li, Lj) = Li, we also know (by tautology) that

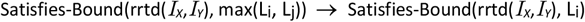

Hence, by the propositional equivalence (P → Q) ^ (P → R) ≡ P → Q ^ R, we know that

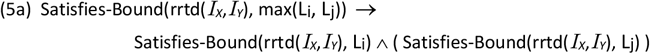

If max(Li, Lj) = Lj, the same arguments hold to show that

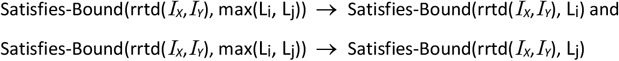

Which are, again, equivalent to (4a), which we have now proven.

Analogous arguments show that, regardless of whether min(Ui, Uj) = Uj or Uj, the following implication holds:

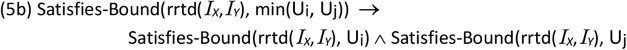

Hence, we can combine (5a) and (5b) to prove (5), because, in general, the following proposition holds:

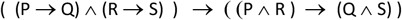

Hence, by proving (4) and (5), we have proven Claim 4.

#### 7.6 Proof of Claim 7: Subsumption Testing of RTRs with Disjunctions (Generalized RTRs)

**Claim 7**: For any two Generalized RTRs, as defined in Section 2.10, GRTR_a_ and GRTR_b_, where

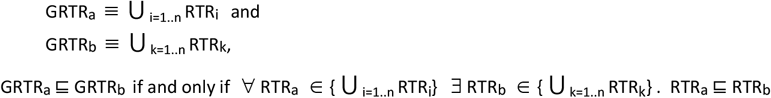

**Proof:** Note that, given the defined semantics of subsumption between pairs of GRTRs and the defined semantics of subsumption between pairs of RTRs, the expression

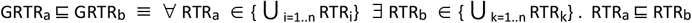

is equivalent to

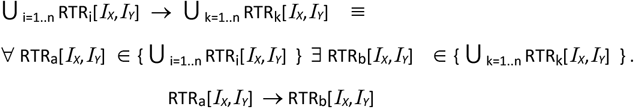

Also note that the righthand side of the logical expression above is equivalent to

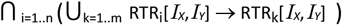

Hence, Claim 6 can be rewritten as

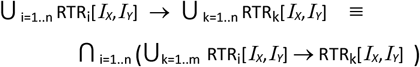

Replacing each RTR predicate in the candidate GRTR and the reference GRTR with the propositional symbols P and Q, respectively, we can simplify the expression to

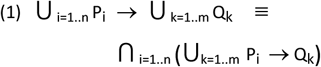

For example,

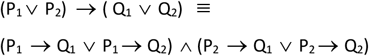

Note how the example is equivalent to the predicate logic expression

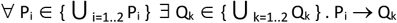

We will prove (1), which is equivalent to Claim 6, by induction.

We begin by proving the simpler assertions (2) and (3) below

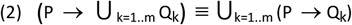

Proof of Inductive step:

We know more generally (2a) R → (S ∨ T) ≡ (R → S) ∨ (R → T) which can be proven by propositional logic:

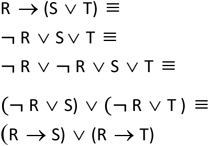

So,

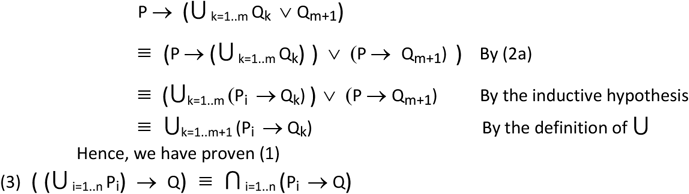

Proof by induction:

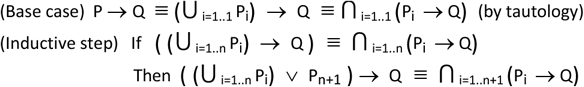

Proof of Inductive step:

We know more generally (3a) (R ∨ S) → T ≡ (R → T) ^ (S → T) which can be proven by propositional logic:

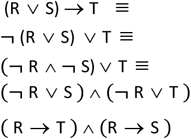

So,

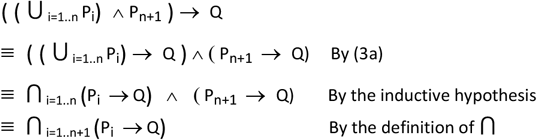

Hence, we have proven (2) Using (2) and (3), we prove (1) by induction.

(Base Case) P → Q ≡ ⋃ _i=1..1_ P_i_ → ⋃ _k=1..1_ Q_k_ ≡ ⋂ _i=1..1_ (⋃_k=1..1_ (P_i_ → Q_k_)) (by tautology) (Inductive step (1a): When adding a proposition to the antecedent disjunction)

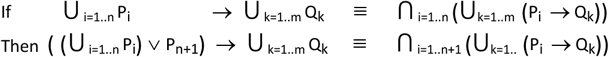

Proof of Inductive step:

Again, we know more generally (2) (R ∨ S) → T ≡ (R → T) ^ (S → T) So,

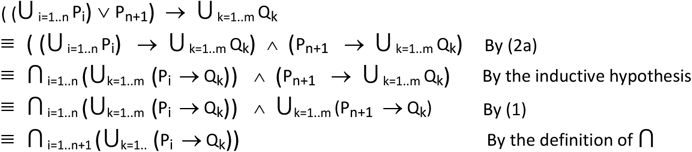

Hence, we have proven inductive step a, i.e., for the case when a proposition is added to the antecedent disjunction.

(Inductive step (1b): When adding a proposition to the consequent disjunction)

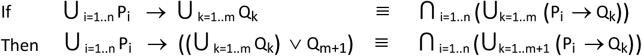

Proof of Inductive step:

Again, we know more generally (1a) R → (S ∨ T) ≡ (R → S) ∨ (R → T) So,

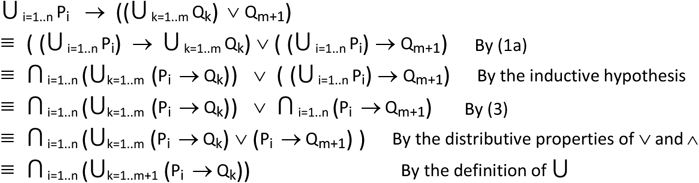

Hence, we have proven inductive step (1b), i.e., for the case when a proposition is added to the consequent disjunction.

Having proven both the inductive steps (1a) and (1b), we have proven (1).

## References

[1] Allen JF. Maintaining knowledge about temporal intervals. Communications of the ACM, 11(26):832–843, November 1983.

[2] Dechter R Meiri I Pearl J. Temporal constraint networks. Artif Intell 1991;49:61–95.

[3] Kautz H, Ladkin P. Integrating Metric and Qualitative Temporal Reasoning. Proc. 9th Nat. Conf. on Artificial Intelligence (1991), pp. 241–246

[4] Meiri I, Combining qualitative and quantitative constraints in temporal reasoning, in: Proceedings AAAI-91, Anaheim CA (1991).

[5] Vilain M, Kautz H. Constraint Propagation Algorithms for Temporal Reasoning. Proceedings of the AAAI Conference on Artificial Intelligence, Volume 5, 1986.

[6] Boland MR, Tu SW, Carini S, Sim I, Weng C. EliXR-TIME: A Temporal Knowledge Representation for Clinical Research Eligibility Criteria. AMIA Jt Summits Transl Sci Proc. 2012;2012:71–80.

[7] Capurro D, Barbe M, Daza C, María JS, Trincado J, Gomez I. ClinicalTime: Identification of Patients with Acute Kidney Injury using Temporal Abstractions and Temporal Pattern Matching. AMIA Jt Summits Transl Sci Proc. 2015 Mar 25;2015:46–50.

[8] Callahan A, Polony V, Posada JD, Banda JM, Gombar S, Shah NH. ACE: the Advanced Cohort Engine for searching longitudinal patient records. J Am Med Inform Assoc. 2021 Jul 14;28(7):1468–1479.

[9] Starren J, et. al. Encoding a post-operative coronary artery bypass surgery care plan in the Arden syntax. Computers in Biology and Medicine. 24(5):411–417. 1994.

[10] CMS Electronic Clinical Quality Measures, 2003. https://ecqi.healthit.gov/ep-ec?qt-tabs_ep=1&globalyearfilter=2023&global_measure_group=3716. Accessed 11/16/23)

[11] Schulz S, Markó K, Suntisrivaraporn B. Formal representation of complex SNOMED CT expressions. BMC Med Inform Decis Mak. 2008 Oct 27;8 Suppl 1(Suppl 1):S9.

[12] Kohane IS. Temporal Reasoning in Medical Expert Systems. MIT/LCS/TR-389. MIT Laboratory for Computer Science. 1986. (http://groups.csail.mit.edu/medg/ftp/kohane/MIT-LCS-TR-389.pdf Accessed 2/27/2023)

[13] Das AK and Musen MA. (1994). A Temporal Query System for Protocol-Directed Decision Support, Methods of Information in Medicine, 33(4), 358–370.

[14] Das AK. Temporal Mediation of Relational Databases for Clinical Decision Support. Doctoral Dissertation, Medical Information Sciences Program, Stanford University. 2002.

[15] Shahar Y. A Knowledge-Based Method for Temporal Abstraction of Clinical Data. Doctoral Dissertation, Medical Information Sciences Program, Stanford University. 1994.

[16] Shahar Y, Musen MA. Knowledge-based temporal abstraction in clinical domains. Artif Intell Med. 1996 Jul;8(3):267–98.

[17] Post AR, Harrison JH Jr. PROTEMPA: a method for specifying and identifying temporal sequences in retrospective data for patient selection. J Am Med Inform Assoc. 2007 Sep-Oct;14(5):674–83.

[18] Capurro D. Secondary Use of Electronic Clinical Data: Barriers, Facilitators and a Proposed Solution. Doctoral Dissertation, University of Washington. 2012.

[19] Hripcsak G, Zhou L, Parsons S, Das AK, Johnson SB. Modeling electronic discharge summaries as a simple temporal constraint satisfaction problem. J Am Med Inform Assoc. 2005 Jan-Feb;12(1):55–63.

[20] El-Sappagh S, Franda F, Ali F, Kwak KS. SNOMED CT standard ontology based on the ontology for general medical science. BMC Med Inform Decis Mak. 2018 Aug 31;18(1):76.

[21] Baader, F., Brandt, S., Lutz, C.: Pushing the EL Envelope. In: IJCAI, Professional Book Center (2005) 364–369

[22] https://en.wikipedia.org/wiki/Disjunctive_normal_form (Accessed 3/1/2023)

